# Contact Tracing Can Explain Counter-Intuitive COVID-19 Trajectories, Mitigate Disease Transmission and Provide an Early Warning Indicator – A Mathematical Modeling Study

**DOI:** 10.1101/2021.09.27.21264174

**Authors:** B Shayak, Mohit M Sharma

## Abstract

The COVID-19 trajectories worldwide have shown several surprising features which are outside the purview of classical epidemiological models. These include (*a*) almost constant and low daily case rates over extended periods of time, (*b*) sudden waves emerging from the above solution despite no or minimal change in the level of non-pharmaceutical interventions (NPI), and (*c*) reduction or flattening of case counts even after relaxation of NPI. To explain these phenomena, we add contact tracing to our recently developed cluster seeding and transmission (CST) model, which is predicated on heterogeneous rather than homogeneous mixing of people in society. With this addition, we find no fewer than four effects which make prediction of epidemic trajectories uncertain. These are (*a*) cryptogenic instability, where a small increase in population-averaged contact rate causes a large increase in cases, (*b*) critical mass effect, where a wave can manifest after weeks of quiescence with no change in parameter values, (*c*) knife-edge effect, where a small change in parameter across a critical value can cause a huge change in the response of the system, and (*d*) hysteresis effect, where the timing and not just the strength of a particular NPI determines the subsequent evolution of the epidemic. Despite these effects however, it is a robust conclusion that a good contact tracing program can effectively substitute for much more invasive measures. We further find that the contact tracing capacity ratio – a metric of the stress to which the tracers are subject – can act as a reliable early warning indicator of an imminent epidemic wave. Extensive simulations demonstrate that whenever there is a drop in capacity ratio during a period of low daily infections, there is a very high probability of the case counts rising significantly in the immediate future.

**Author summary:** Close to two years into the pandemic, the trajectories of COVID-19 in different places and at different times have shown wild variations and confounded modeling and forecasting efforts. Our new mathematical model can help to explain these variations. Some solutions of our model are non-standard but realistic. For example, we find an epidemic curve where daily cases remain on a plateau for a long time before suddenly exploding into a wave, despite interventions remaining constant throughout. We also find solutions showing that a specific intervention, for example capacity reduction at public gatherings, is very effective if implemented early on in a wave but useless if implemented a little later. Our proposed early warning indicator can be a game-changer for epidemic forecasting and model-based intervention strategies. Current forecasting algorithms have the weakest performance at the inflection points where there is an abrupt change in trend in the daily infection rates. The early warning indicator can give us advance notice of an approaching inflection point, and enable the authorities to take preventive measures before a wave actually arrives. Our results indicate that close communication between contact tracing personnel and public health authorities can achieve synergistic mitigation of the pandemic.

## INTRODUCTION

### §1. Background

The dynamics of COVID-19 trajectories in different countries have shown features which are unexpected from a classical epidemiological viewpoint. “Plateaus” or extended periods of low and nearly constant daily case rate have been seen for example in USA (June 2021), UK (April to May 2021), India (January to March 2021), Australia (November 2020 to June 2021) Canada (June to August 2020), Germany (May to August 2020), Uruguay (March to November 2020) and Taiwan (March 2020 to April 2021). In each case, the quiescent period was shattered by a sudden wave; in at least some instances, for example India and Taiwan, non-pharmaceutical interventions (NPI) had been relaxed long before the wave arose and there was no change in public health policy immediately preceding the wave. In UK, a wave ostensibly driven by the extreme transmissibility of the B1.617.2 (“delta”) variant began in late May 2021; despite this, all restrictions were relaxed on 19 July 2021, after which the case counts went down. A senior scientist with huge COVID-19 modeling experience admitted to being flummoxed by this development [1]. In India also, following the horrific second wave, B1.617.2 (the dominant strain) currently remains under control. In USA however, the cases have been following a “roller-coaster” trend, which has left epidemiologists confused [2, 3]. In the past two weeks, waves began again in many European countries, leading to reimposition of social restrictions after many months. The prevalence of different mutants with time in USA [4] and India [5] is also interesting. In both these countries, B1.617.2 has become dominant during the most recent wave. In each case however, wildtype B1 (usually treated as “other”) has clung to a steady, small but non-zero fraction of total infections while the apparently more transmissible [6] B1.1.7 (“alpha”) variant has been decimated to nearly zero prevalence.

Mathematical models for COVID-19 spread are of several types, the two most widely used being compartmental or lumped parameter models [7–13] and agent-based models [14–19]. So far, no compartmental model has succeeded in explaining any of the above ‘anomalies’. While agent-based models are in theory capable of generating or predicting any epidemic trajectory, a convincing, comprehensive explanation of the counter-intuitive disease transmission patterns mentioned above remains difficult to find. A handful of modeling studies so far [20–23] have found the plateau as a non-marginal solution, without accounting for a sudden wave (a rebuttal [24] exists for one of these analyses). Another few studies [25–27] have found the sudden waves. In the first of these, the successive waves are progressively smaller in size, so its real world accuracy is yet to be explored. The second relies on certain ansatzes. The third is the most realistic and agrees well with many of our findings, but it does not feature a plateau. Finally, there are some network models of generic epidemics [28–31], which are yet to achieve significant success with COVID-19.

Since a sudden wave can have disastrous consequences for healthcare systems, the question arises as to whether it can be predicted in advance. Drawing reference to the critical slowing down phenomenon [32], Dablander et. al. [33] have presented a detailed discussion on this point. These authors consider a compartmental model with time-dependent parameters and show that many commonly proposed indicators actually decrease rather than increasing before a wave, and are hence useless. This issue has been further highlighted in a recent meta-analysis [34] of the performance of various COVID-19 forecasting algorithms in Germany and Poland. This analysis found that the maximum prediction error occurred at the inflection points where there was a change in trend in daily case rates i.e. a transition from plateau to wave or from increasing to decreasing. Ironically, from the public health perspective, it is these very points which are of the greatest significance.

In a recent work, published as Ref. [35], we have proposed a new infectious disease dynamical model called cluster seeding and transmission (CST) model. The underlying principle of this model is that human interaction is not homogeneous but heterogeneous – in particular, unmasked interactions at little or no separation, which are most liable to spread COVID-19, remain primarily confined within small and closed groups of people called clusters. Reference [35] can account for the plateau solution as well as a sudden transition to wave, but beyond that, its descriptive power is still limited. Here we have added contact tracing to this model, as described in Methods later in the Article. We now demonstrate that with this addition, the CST model can explain all the anomalies mentioned above as well as provide a potential early warning indicator.

## RESULTS AND DISCUSSION

### §2. Complex effects in the model

Two preliminary considerations : we assume (*a*) no reinfection and (*b*) no vaccination. A detailed analysis of temporary immunity as well as vaccination is appropriate for a future, separate study; a very approximate estimate of the consequences of vaccination can be obtained simply by considering a fraction (vaccinated percentage times vaccine efficacy) of the population to be pre-immune.

In CST model, a person’s cluster consists of close contacts such as workplace colleagues with whom s/he regularly participates in potentially risky activities such as going to the movies and dining outdoors. Reference [35] has identified that socializing external to cluster (SEC) events like wedding and birthday parties, where people intentionally mix outside their cluster, are very important transmission venues (these events are conceptually similar to the transient simplices of Ref. [36] although the underlying network structures are different in the two models). The number *n_S_* of people participating daily in SEC events is a fundamental parameter in the model. Reference [35] has shown that SEC events, while adding a small contribution to the population-averaged contact rate, can make a gigantic contribution to the caseload. We have called this the *cryptogenic instability*, the latter word referring to the destabilization of the plateau solution into a wave. This is one of the four non-classical effects present in our model. The other three are the novel results of this Article.

The second effect is *critical mass effect*. This refers to the fact that, for certain choices of parameter value, the disease can simmer for a long time before suddenly exploding into a wave. We show an example trajectory in Fig. 1 below, with *n_S_* = 1800 and the other parameter values being given in Methods. All simulation results are for a model city of population 3,02,400.

**Figure 1 :**
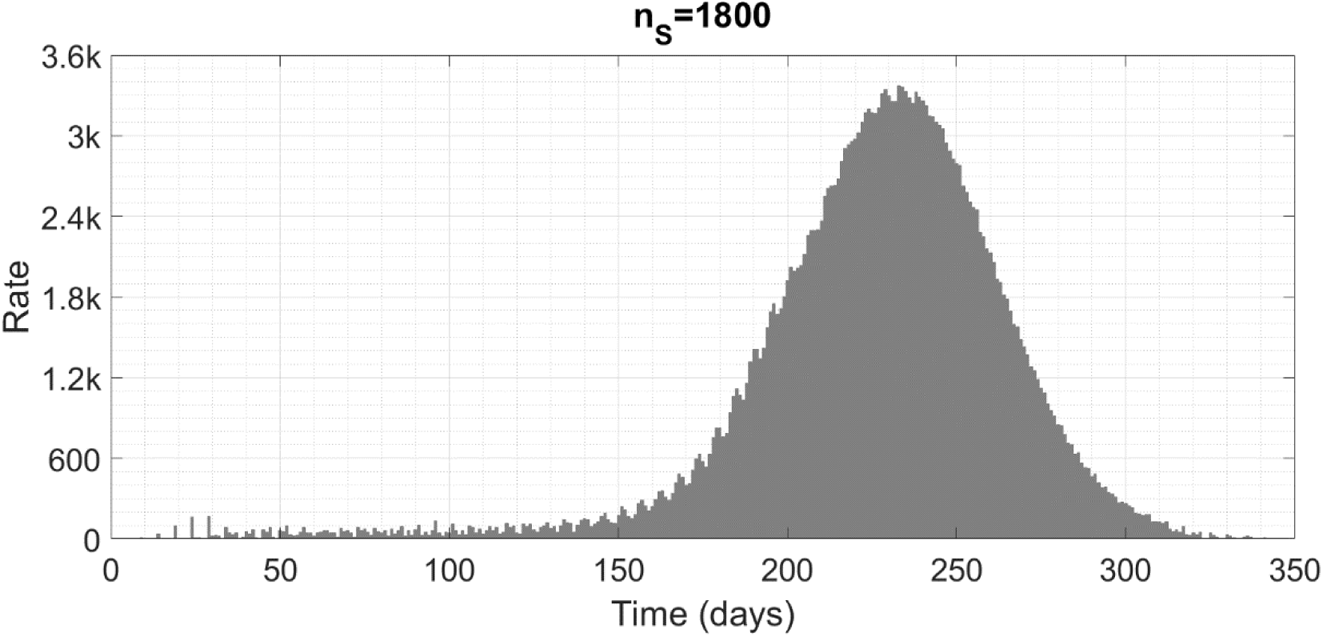
Case trajectories showing the critical mass effect. The numbers of new cases emerging per day are shown as grey bars. The symbol ‘k’ denotes thousand.

We highlight that this entire trajectory – with a nearly constant case rate from 40 to 130 days and a huge wave after 150 days – has been generated with parameters kept invariant throughout.

The third effect is *knife-edge effect*. This refers to the fact that if we vary one parameter while keeping all others fixed, and ask for the cumulative caseload at the end of the outbreak as a function of the varying parameter, then we do not see a gradual change from a low to a high caseload. Instead, there is a sudden jump from containment (plateau) to herd immunity (wave) over a very narrow range of the parameter value – on either side of the jump, the caseload remains almost constant as the parameter is varied. In Fig. 2 below, we show the final size and total duration of the epidemic obtained by varying *n_S_*. The eventual cumulative caseload is depicted a blue line attaching to the left hand *y*-axis and the epidemic duration as a green line attaching to the right hand *y*-axis. We show similar plots for variation of other parameters in the Supporting Information (SI).

**Figure 2 :**
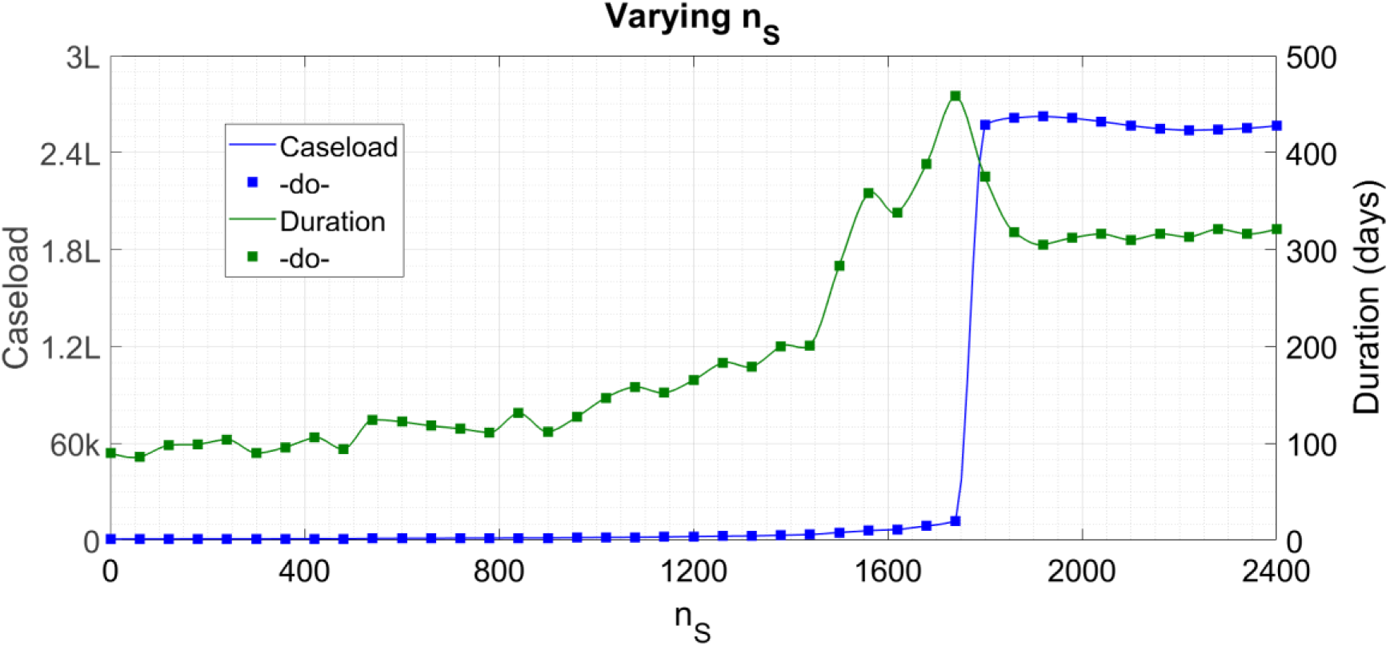
Cumulative caseload (blue) and duration (green) of the epidemic as n_S_ is varied and all other parameters held to their default values. The symbol ‘k’ denotes thousand and ‘L’ hundred thousand.

The final effect is *hysteresis effect*, in which the past behaviour of the epidemic influences its present evolution. For example, with other parameters same as in Fig. 1, if the epidemic evolution starts with *n_S_* = 1500 then we see a plateau while if it starts with *n_S_* = 1900 then we see a wave (check from Fig. 2 that the knife-edge lies at approximately *n_S_* = 1750). However, if a wave is already initiated at 1900, then a reduction to 1500 after 100 days has no effect on it whatsoever. A much more drastic reduction – to *n_S_* = 300 for instance – is required to stop the wave in its tracks. Conversely, relaxing a lockdown may or may not cause a rebound wave depending on the daily case rate at the time of relaxation. Similarly, an augmentation of contact tracing capacity in response to a wave may prove highly effective if implemented early enough but much less effective if implemented later. In Fig. 3 below, we show the time trace for the second of these two situations. The starting *n_S_* is 1900, lockdown is implemented by reduction to 300 and unlock by relaxation to 1500. The daily new cases are grey bars as in Fig. 1 and the cumulative caseload is a blue line attaching to the right hand *y*-axis.

**Figure 3 :**
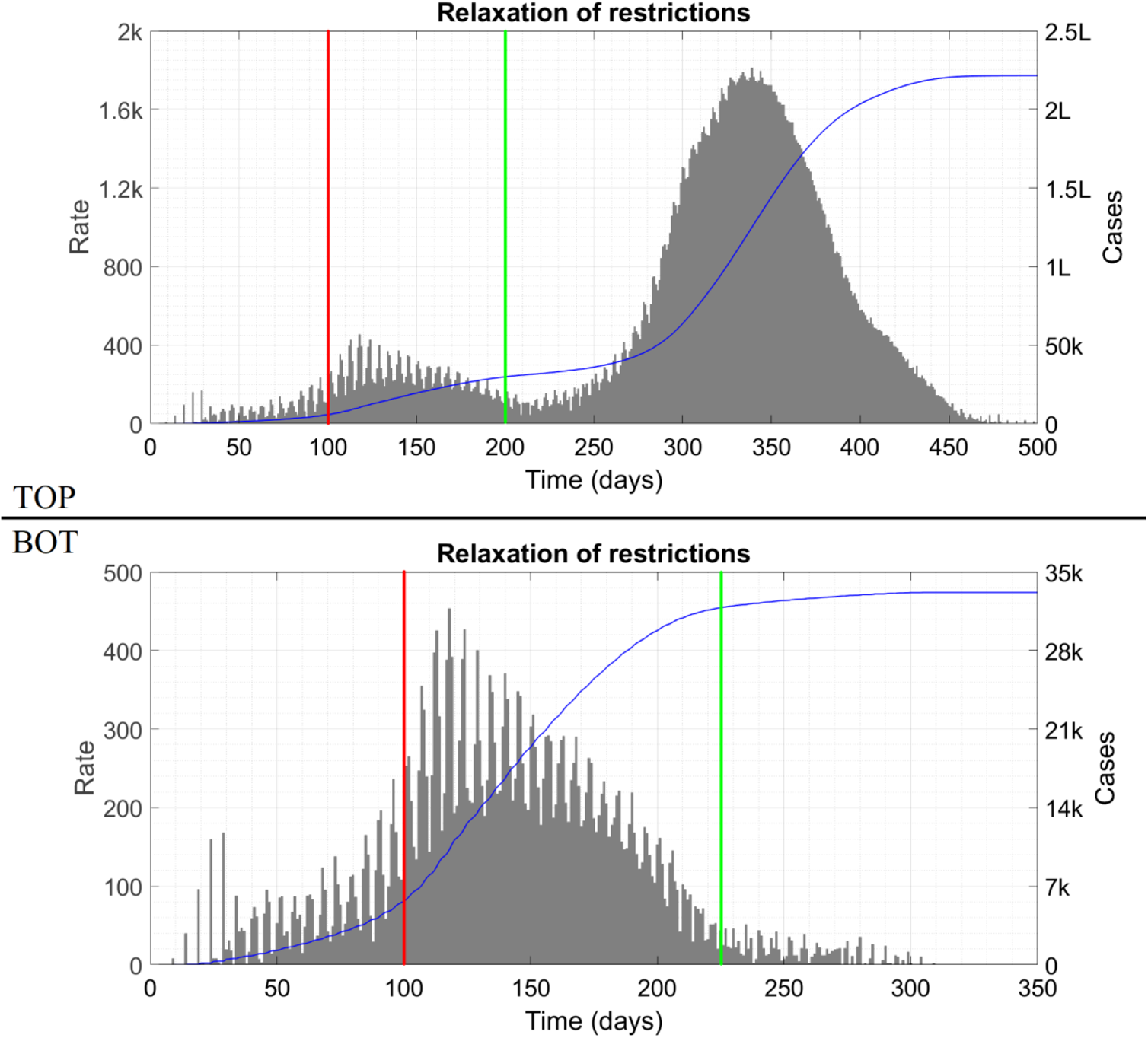
Top panel shows the effects of increasing n_S_ from 300 to 1500 on day #200 while bottom panel shows the effects of the same change implemented on day #225. In both cases, the lockdown is applied on day #100, indicated by the red vertical line. Note that upto day #200, both trajectories are identical – they appear different only because of the axis scalings. The unlock is indicated by the green vertical line. The symbol ‘k’ denotes thousand and ‘L’ hundred thousand.

We show the time traces for the other situations in SI. Figure 3 as well as the supplementary figures show that, as in any classical hysteretic system, the epidemic trajectory at any instant is influenced not only by the parameter values prevailing at that instant but also by the trajectory itself at past times.

### §3. Analysis of the effects

While critical mass effect in the CST model without contact tracing has been identified in Ref. [35], the tracing is responsible for enhancing it substantially. In the absence of tracing, for the same parameter values we can get either an isolated handful of cases (for a small initial condition) or a full-blown wave (for a larger initial condition) but not both in series. With the tracing included however, Fig. 1 shows just such a solution, and this is a hallmark of COVID-19 trajectories round the world. We can also see that critical mass effect can result in a wave occurring months rather than days after a NPI relaxation, when nobody is expecting trouble, as happened in India, Taiwan and elsewhere.

The qualitative reasoning behind the solution of Fig. 1 is as follows. Firstly, we note that contact tracing dramatically reduces the number of at large cases. With the tracing probabilities set to their Fig. 1 values (see Methods), excluding the chance that a 24-member cluster is caught at seeding itself, there is a 24 percent probability that the cluster develops just one at large case and another 21 percent probability that it develops four at large cases. Now, in Fig. 1, the parameter values happen to be chosen in such a way as to generate a plateau if the contact tracers can access every emerging symptomatic case (note that the ‘if’ condition is not actually satisfied by the situation at hand). After the seeding period (first 40 days), the number of fresh cases occurring per day is such as to stretch the tracing infrastructure to its limit or very slightly beyond. Thus, for a long time, nearly all feasible cases are traced and the plateau continues with a very slight increasing trend. Now, this plateau is not perfectly smooth but consists of fluctuations about an almost constant envelope. The increasing trend means that, as time goes on, days of locally high cases seed more clusters than the tracers can access and isolate. These untraced clusters in turn start supplying considerable numbers of at large cases. And now we enter a vicious circle, for more at large cases with constant contact tracing capacity means yet more untraced clusters and at large cases. This vicious circle manifests as the sudden wave. Thus we can say that the emergence of the wave is the result of a feedback process between the contact tracing and the case trajectory itself. This mechanism is similar to the one proposed in Ref. [27]; in that study however, the trajectory before the sudden wave is also exponential growth, just at a slower rate. Critical mass effect is at best a thorn in the side and at worst a nemesis of public health planners, since, right until the wave occurs, the case counts alone provide scant evidence that it is imminent and that an increase in NPI or compliance therewith is urgently called for.

Like the critical mass effect, knife-edge effect confounds the efforts of epidemic predictors and planners. The risk here is that the transition from tractable (plateau) to intractable (wave) behaviour is very sharp, and the system response on either side of the knife-edge is qualitatively the same. Hence, if a region is operating in plateau mode with a certain level of NPI, it is impossible to guess how much further NPI relaxation is possible without the plateau being destabilized. Incremental relaxation – a policy used almost worldwide during the initial exit from lockdown – will scarcely produce results until the knife-edge is hit, when the consequences will suddenly become drastic.

As an example of hysteresis effect, we consider the state of Maharashtra in India, including its two primary case-driver cities Mumbai and Pune. During January and February 2021, cases in these cities were constant or decreasing even though NPI were not very stringent. When a wave arose in early March, the reproduction number *R* was relatively low [37] and the State Government initially tried a less invasive intervention approach such as imposing night curfews and increasing mask compliance. As per compartmental epidemic models, this alone should have worked. If *R* was below unity in January at a certain level of NPI, then reverting to that level of NPI in March should have brought it below unity again and stemmed the growth of cases. This however was not what happened – the daily case counts grew relentlessly and hospitals became overwhelmed, to the distress of authorities and the public alike. Only when a full lockdown was imposed did the numbers start going down again. Hysteresis effect can explain this observation completely. The public health implication of this effect is that shorter periods of strong interventions are superior to longer periods of weaker interventions at combating a rise in cases.

All of these effects combine together to explain the bewildering variety of COVID-19 trajectories seen all over the world. Anomalously high case counts, as in USA and Israel, might indicate the crossing of a knife-edge during the relaxation process – thereafter, hysteresis effect ensures that the trajectories do not respond to mild NPI. Unexpectedly low counts, for example in India and UK [1], might indicate operation on the lucky side of a knife-edge. Roller-coaster effects, as occurring in USA, might be related to changes in SEC events. For example, a rise in cases might cause more people to forgo attendance at parties and bashes, and hence automatically bring *n_S_* down to a level where cases begin falling (see also Ref. [23]). Since the SEC events have a small contribution to total interaction and are currently not accorded special status as per public health policy, fluctuations in *n_S_* will not show up in mobility analyses, and the changes in disease trend will appear mystifying.

The complex effects underlying corona trajectories might also influence analyses of transmissibility of mutants. For example, the transmissibility of B1.1.7 variant appears to have been overestimated since it was eliminated in competition while B1 survived. Dominance of B1.617.2 relative to other variants will also be facilitated if the serial interval for this variant is lower than for the other variants, which two studies [38, 39] suggest to be the case. The epidemic doubling time *T_d_* during exponential growth is defined as *T_d_* = (log 2)*T_s_* / log *R* where *T_s_* is the serial interval [12]; a smaller *T_s_* results in faster doubling at the same *R*.

### §4. Early warning indicator

So far we have focused on the variability of COVID-19 trajectories, both in reality and in our model. We now ask, is there method in this madnessThat is, we want to know whether everything is upto chance or whether it might be possible to accurately perform at least short-term forecasts of COVID-19 trajectories, in particular of the sudden waves. Quantitative predictions based on large numbers of variables and parameters are at risk of being inaccurate due to incorrect parameter estimates. Hence we seek a single, physically significant quantity which might cast insight into the COVID-19 dynamics as a whole. Such a quantity can act as an early warning indicator.

To get this quantity, we need to introduce two numbers related to contact tracing. The first is *n_CT_*, the number of contacts who ought to be traced per day if the tracers operate as per their usual protocols. This will be dependent on the number of new cases emerging per day and hence will be a function of time. The second number is *CT*_max_, the maximum number of contacts which the city authorities can trace in one day. This will depend on the infrastructure and personnel available to the city, and will be more or less constant over short periods. Define the *capacity ratio ρ* as *ρ* = *CT*_max_/*n_CT_* if *CT*_max_ < *n_CT_* and *ρ* = 1 otherwise. Qualitatively, *ρ* measures the stress on the contact tracing system. When the absolute case counts are low, *ρ* will equal unity but when the counts are high, this will not hold true as the tracers will have to let go of some cases due to resource constraints.

In our analysis we have found that if *ρ* becomes less than unity during a quiescent phase, then there is a very real threat of a wave. While *ρ* = 1, all feasible contacts are getting traced, i.e. the number of potential contacts emerging per day is less than *CT*_max_. This automatically puts a cap on the maximum number of daily cases and precludes a wave. On the other hand, when *ρ* decreases below unity, there is a surplus of at large cases which initiates the vicious feedback circle between cases and untraced clusters. We expect that, during the first few days at least, the surplus will be small – it will take some time for the untraced cases to be amplified to a level where the growth becomes uncontained. During this time, the actual case counts will remain small but *ρ* will dip below unity, thus acting as an early warning indicator.

To verify our hypothesis, we perform extensive simulations. As a representative example to include here, we consider Fig. 1. In this figure, the parameter value *n_S_* = 1800 generated a huge wave after a faux plateau; if we reduce *n_S_* to 1750 then we get the plateau only (again recall the knife-edge from Fig. 2). In Fig. 4 below we show the epidemic history during the initial 120 days for these two values of *n_S_*. We also show *ρ* as a blue line attaching to the right hand *y*-axis.

**Figure 4 :**
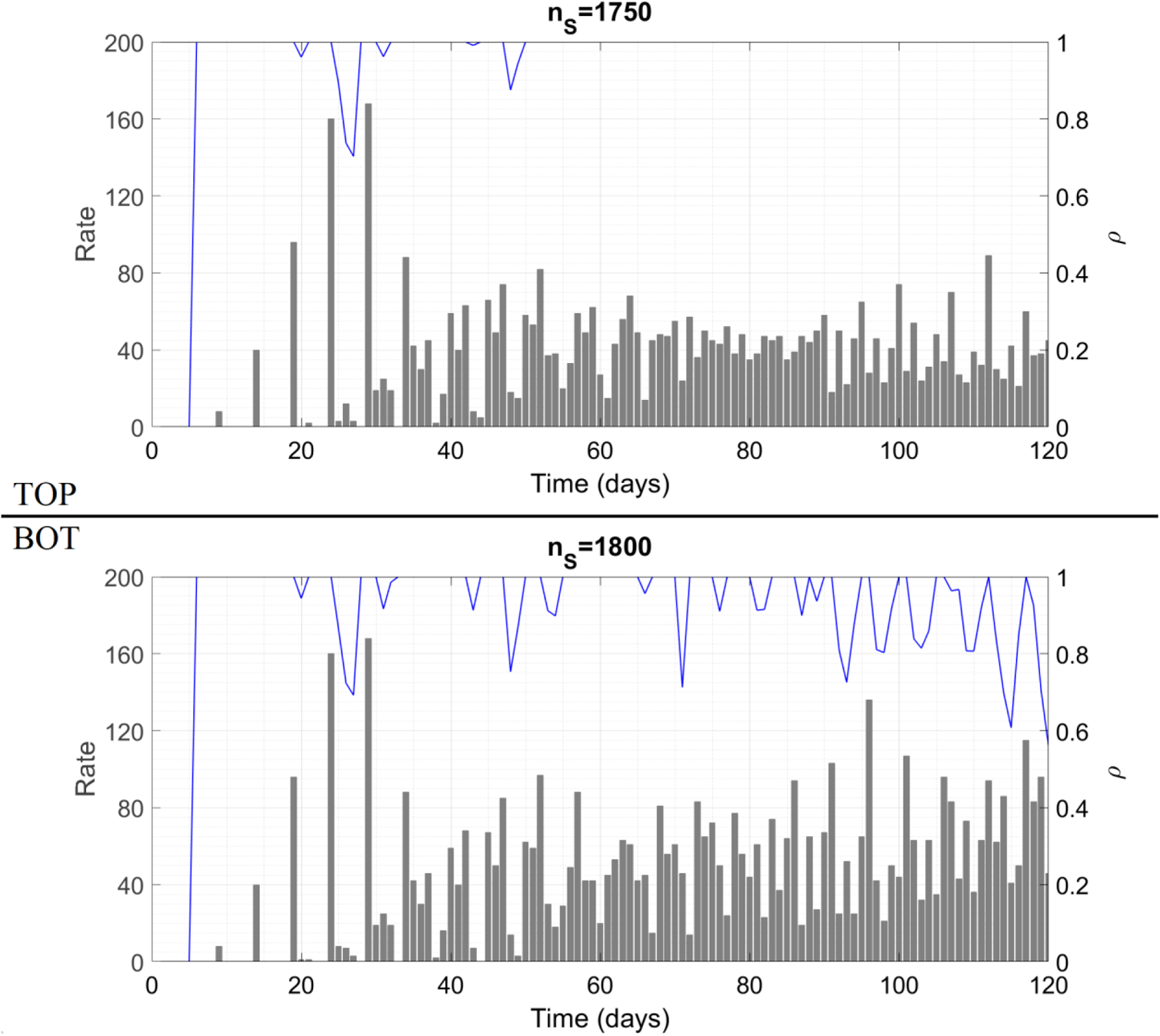
Case trajectories and capacity ratio as a function of time during the first 120 days for two different values of n_S_. After 120 days, the top panel continues as is while the bottom panel is the same as Fig. 1.

Indeed, the case trajectories in the two panels look nearly identical. But while *ρ* for the top panel is unity almost throughout, *ρ* for the bottom panel shows marked deviations which increase as time goes on. The bottom panel does show a very slight increasing trend superposed on the plateau but this can easily be missed or taken for an unimportant fluctuation. The fact that the absolute case counts are higher in the bottom panel than in the top is insignificant since a plateau is a plateau, and absolute case counts have no meaning. But the trend in *ρ* in the two plots is very different, and this difference is crucial as the subsequent evolution demonstrates (the final case counts in the two situations differ by a factor of more than 12).

In SI, we present many more time traces of *ρ* in borderline situations, for different NPI measures and parameter values. The conclusion which all the plots support is that *ρ* performs well as an early warning indicator in each case.

### §5. Conclusion and policy implications

In our CST model of epidemic propagation with contact tracing, we have identified four non-classical effects which can help to explain the perplexing variety of COVID-19 trajectories seen worldwide. We have also shown that the capacity ratio *ρ* might act as an early warning indicator of an imminent wave. In Methods, we present a detailed discussion of the limitations of the model and their consequences. Many of the limitations arise from assumptions needed to make the model deterministic rather than fully-agent based. When these assumptions are relaxed, the cryptogenic instability, critical mass and hysteresis effects are expected to remain as they are while the transition from containment to herd immunity in the knife-edge effect is expected to become more gradual. Despite these limitations, CST model with contact tracing appears to be superior to other models at explaining and predicting COVID-19 epidemiological curves.

We now briefly discuss the public health policy implications of our work. Policy decisions must be formulated on the basis of results which remain qualitatively unaffected by changes in parameter values or refinements in the model structure. The first such result is that SEC events activate the cryptogenic instability and can cause a high caseload. For this reason, size limits on such gatherings must be imposed and enforced. Public health messaging may focus on narrowing the scope of one’s social interactions as a substitute to reducing their frequency. Testing of asymptomatic persons, an intervention not considered here, may turn out to be most effective if implemented on the prospective attendees immediately prior to SEC events; we shall analyse this in detail in the future.

The second robust conclusion is that contact tracing is of great importance in combating the spread of COVID-19. To facilitate the contact tracers’ job, people may be encouraged to keep private records of every person with whom they have had close, unmasked interactions. Then, in case a person turns out corona positive, the entries for the last five or seven days in his/her record may be used to quickly access potential contacts and also trace the case’s SEC transmissions. Compared with mobile phone-based contact tracing methods [40], this technique has two benefits : (*a*) the electronic methods do not differentiate between an interaction which took place with full COVID-appropriate behaviour and one which did not, and (*b*) there is no question of the personal records being used for surveillance without one’s knowledge. Two cautionary points regarding an effective contact tracing program are given in SI.

The third conclusion which appears to be robust is that saturation of the contact tracing infrastructure indicates the imminent arrival of a wave. When such saturation is observed, a rollback of reopening measures should be announced immediately and maintained until the strain on the contact tracers is eased. Augmenting the contact tracing capacity by hiring professional contact tracers will enable further reopening, and, in concert with vaccination, pave the way towards possible elimination of the disease in the longer term. We shall consider such phenomena in our future studies.

## METHODS

### §6. Cluster seeding and transmission (CST) model – prior work

We first give a brief summary of the basic CST model presented in Ref. [35]. An unstated or at best understated assumption at the heart of all lumped parameter or compartmental models is that of homogeneous mixing – the assumption that any random person has equal probability of interacting with, and hence transmitting the virus to, any other random person taken from the entire population. In our cluster-based model we have gone with the more realistic assumption that a person is more likely (perhaps by order of magnitude) to interact with (and consequently infect) family members and friends rather than strangers. We have expressed this by dividing the population into clusters or small groups of people with dense links among each other and few or no links outside. Intra-cluster transmission of virus is certain and rapid. Inter-cluster transmission is rarer and (as per the model assumptions) occurs in two ways. These are unintentional cluster transition or UCT events which involve accidental transmission in public places, and socializing external to cluster or SEC events where people from different clusters intentionally get together. By calculating the various transmission probabilities, we have arrived at a computational model for the spread of the disease.

We now give a brief description of CST model. Time is discrete and is measured in days. The variables and parameters are as in Table 1 below (this table is adapted from Ref. [35]). The default values are those used in Ref. [35] to generate a plateau solution, and carry over to this Article with a few exceptions to be mentioned as appropriate. Since we assume uniform mixing among clusters (rather than individuals), the domain of validity of the model is a city rather than a state or a country. This is important – the model is not applicable to a whole state or country at once. To get the curves for such a larger region, we must break it up into cities and explicitly account for travel among the cities. This restriction on the domain size applies to any model where mixing is heterogeneous – homogeneously mixed models allow greater flexibility but at the cost of descriptive and predictive accuracy, as discussed in §1.

**Table 1 :** Variables and parameters in the basic CST model. Parts of this Table are reproduced verbatim from Ref. [35] (our own work). We do not assign a default value to n_S_ (it was zero in Ref. [35]) since it is varied continually for the simulation results.

| Variable | Significance |
| --- | --- |
| $y_i$ | Cumulative number of socially active cases upto and excluding day $\#i$ |
| $\Delta y_i$ | Number of new socially active cases cropping up on day $\#i$ |
| $z_i$ | Cumulative number of insusceptible clusters upto and excluding day $\#i$ |
| $\Delta z_i$ | Number of new clusters turning insusceptible on day $\#i$ |
| $f_i$ | Cumulative number of cautious household cases upto and excluding day $\#i$ |
| $\Delta f_i$ | Number of new cautious household cases cropping up on day $\#i$ |

| Parameter | Significance | Default value |
| --- | --- | --- |
| $N$ | Total population | 3,02,400 |
| $h$ | Household size | 3 |
| $N_1$ | Socially active population | 1,00,800 |
| $N_c$ | Number of clusters | 4200 |
| $s$ | Size of the cluster | 24 |
| — | Cluster sequence | [1; 3; 6; 7; 5; 1] |
| $T_s$ | Serial interval | 5 days |
| $n_U$ | Number of people participating daily in UCT events | 10,000 |
| $P_U$ | Probability of transmission at a UCT event | 0.15 |
| $n_s$ | Number of people participating daily in SEC events | — |
| $m_s$ | Number of people infected by one case at a SEC event | 2 |
| — | Time between exposure and start of transmissibility for all cases | 5 days |
| — | Duration for which all cases transmit the disease to others | 3 days |

We count a person as a case on the day s/he first turns transmissible. Considering a city of total *N* people, we have partioned them into *N*_1_ ‘socially active’ people who can contract the disease on their own and *N* − *N*_1_ ‘householders’ who can contract the disease only from a socially active individual. We have then divided the *N*_1_ active people into *N_C_* clusters of equal size *s*. When a cluster is seeded, i.e. when the first member of a cluster with all susceptible persons is exposed to an infective dose of the pathogen, we assume that cases crop up in the cluster over the next few days following a fixed, definite sequence. The *j*^th^ element of this sequence denotes the number of new cases cropping up in the cluster *j* serial intervals following seeding (for this purpose, the serial interval must be taken as an integer number of days). This sequence approximately captures the reproduction number *R*_0_ of the disease (*R* is the number of secondary transmissions from one infected individual, and *R*_0_ is its initial value when everyone is susceptible) and the serial interval (the time between primary and secondary case). The last two rows of Table 1 indicate our assumption that all cases turn transmissible 5 days following exposure and remain transmissible and at large for 3 days (after this duration, asymptomatic cases recover while symptomatic ones show their symptoms and go into quarantine). We shall in general continue using the same values here. For more details of the prior work, we must request that you consult Ref. [35]; we now proceed to the novel contributions of the present Article.

### §7. Implementation of contact tracing

Contact tracing refers to the process in which, when a case is detected, the people with whom s/he has interacted recently are identified and recommended to isolate for a few days so as to prevent further spread of infection. Here we make two fundamental assumptions regarding the tracing process :

- There is forward contact tracing only, starting from symptomatic cases. This means that when a symptomatic case with unknown source of exposure tests positive, the authorities attempt to identify all of his/her potential secondary cases, but do not attempt to identify the source of exposure and other secondary contacts thereof. We also assume that there is no random testing of asymptomatic persons.
- The process timings involved are such as to ensure that cases who are successfully caught by the contact tracers are sent into quarantine during their non-transmissible incubation period, i.e. traced cases transmit the disease to no one else.

A toy example will help to clarify the implications of the above assumptions. For this example we ignore the cluster structure of the population. Let us say Alfa is an asymptomatic case who transmits the virus to Bravo and Charlie on day #0 (as mentioned in Ref. [35], we use the ICAO letter codenames to denote random persons – overlap with names of viral variants is purely accidental). On day #5, both of them turn transmissible – Bravo transmits to Delta, and Charlie to Echo and Foxtrot, all on the same day. Bravo remains asymptomatic while on day #8, Charlie turns symptomatic, reports to the authorities and tests positive. By the first assumption, Echo and Foxtrot are rounded up by the contract tracers but there is no attempt to track down Alfa. As a result of this lapse Bravo remains undetected as well, and so does Delta. Charlie’s secondary cases Echo and Foxtrot turn transmissible on day #10; the second assumption implies that they are successfully captured within day #9. This discussion might appear a little unwieldy so we show the transmission sequence in a schematic form in Fig. 5 below.

**Figure 5 :**
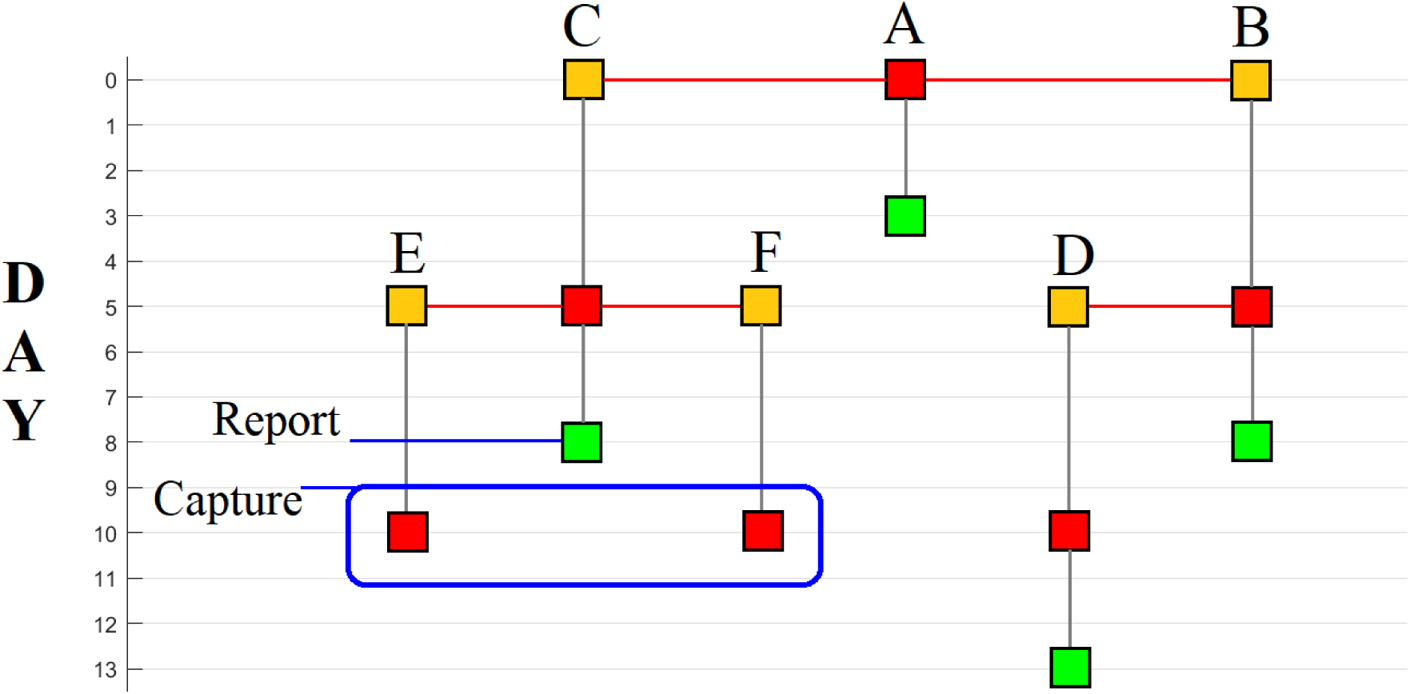
Timeline of different cases in the hypothetical example to demonstrate the contact tracing assumptions. Squares denote individual persons. Yellow means exposed but not transmissible, red means transmissible while green means recovered. The grey vertical lines indicate transition of one person from one state to another while red horizontal lines denote transmission of the disease from person to person. The 5- and 3-day intervals between exposure and infectiousness, and infectiousness and recovery are consistent with the basic CST model assumptions. Note how the two assumptions regarding contact tracing result in capture of Echo and Foxtrot before they turn transmissible while failing to trace the Bravo-Delta branch of the transmission.

Thus we can see that the two assumptions introduce errors in opposite senses. A more sophisticated contact tracing system can achieve backward tracing (catching Alfa from Charlie and hence Bravo and Delta from Alfa) while a slowly functioning system with delays in testing etc can result in secondary cases not being caught before starting spread.

The second assumption allows us to define exactly two classes of cases – at large cases, who spend three full days spreading virus just as in Ref. [35] and quarantined cases who spend zero days infecting others. We now account for the presence of social clusters in a realistic manner. Let *P*_0_ be the probability that a random case is symptomatic, and *Q*_0_ = 1 − *P*_0_ the probability that s/he is asymptomatic. We assume that when a symptomatic case reports to the authorities, the entire cluster to which s/he belongs is successfully identified with probability *P*_1_ and missed with probability 1 − *P*_1_. Further, UCT and SEC transmissions which this case has caused are identified with probabilities *P*_2_ and *P*_3_ respectively.

We use an example rather than a general theoretical discussion to understand how contact tracing will work on a cluster, noting that the example is very easy to generalize. For this example, we use the cluster sequence [1; 3; 6; 8; 5; 1] which also features in very many of the simulation runs. This sequence is adapted from Table 1, with the reason for the change coming below. All clusters are first seeded via UCT or SEC events; again for definiteness, let us focus on the former. Let the example cluster be named Team, let it consist of persons Alfa through Xray (total 24) and let the at large case Yankee be the one who exposes Alfa to the pathogen in a UCT event. In the absence of contact tracing, the cluster sequence implies that

- Alfa (1 person) is exposed by Yankee on day #0 and turns into a case on day #5
- Bravo, Charlie and Delta (3 more persons) are exposed by Alfa and turn into cases on day #10
- Echo through Juliett (6 more persons) are exposed by one or more of Bravo through Delta and turn into cases on day #15
- Kilo through Romeo (8 more persons) are exposed by one or more of Echo through Juliett and turn into cases on day #20
- Sierra through Whiskey (5 more persons) are exposed by one or more of Kilo through Romeo and turn into cases on day #25
- Xray (1 more person) is exposed by one or more of Sierra through Whiskey and turns into a case on day #30

Now incorporate contract tracing on Team. There is a probability *P*_0_ that Yankee is symptomatic and *Q*_0_ that he is asymptomatic. In the latter case, there is no hope of catching Alfa; in the former case, there is a probability *P*_2_ that Alfa is identified by the contact tracers and sent into quarantine. If this happens, then the only case in Team is Alfa, who is quarantined at the get-go. In such a situation, we classify Team as a cluster of Type 1. The probability that Team, and by extension any other cluster, is of Type 1 is thus *P*_0_*P*_2_.

If Yankee is asymptomatic (probability 1 − *P*_0_) or if he is symptomatic (*P*_0_) but the tracers fail to identify Alfa as his secondary UCT case (1 − *P*_2_), then Alfa becomes an at large case. The probability of this happening is 1 − *P*_0_ + *P*_0_ (1 − *P*_2_) which is 1 − *P*_0_*P*_2_. Then, Alfa transmits the virus to Bravo, Charlie and Delta and also participates in UCT and SEC events. Now, consider the case that Alfa is symptomatic (*P*_0_). If yes, the contact tracers get to work and, with probability *P*_1_, capture the entire cluster Team including Bravo, Charlie and Delta. Assume the complement 1 − *P*_1_ to denote a tracing error or roadblock where the opportunity to catch Team is irreversibly lost. The tracers also capture Alfa’s UCT and SEC transmissions with probabilities *P*_2_ and *P*_3_. Captures of UCT and SEC transmissions by members of Team are however accounted for while implementing contact tracing on the secondary clusters, just as we have factored in Yankee while doing the calculation for Team. Hence, for analysing Team, they do not require our further consideration. Bravo, Charlie and Delta contract the infection in quarantine and further spread of the disease within Team is halted. In this situation, we call Team a cluster of Type 2. The probability of this occurring is (1 − *P*_0_*P*_2_) *P*_0_*P*_1_.

If Alfa is asymptomatic however (*Q*_0_), then Bravo through Delta perforce become at large cases, transmitting the virus to the next level in the cluster i.e. to Echo through Juliett (and also participating in UCT and SEC). Any and all of Bravo through Delta might be symptomatic or asymptomatic – if at least one is symptomatic (1 − *Q*_0_^3^, the complement of the probability that all three are asymptomatic) then Team is grounded at this stage with probability *P*_1_. Echo through Juliett become quarantined cases and we classify Team as Type 3. The probability of this occurring is (1 − *P*_0_*P*_2_) *Q*_0_ (1 − *Q*_0_^3^) *P*_1_.

Similarly, if everyone upto Delta is asymptomatic (*Q*_0_^4^) but at least one among Echo through Juliett is symptomatic (1 − *Q*_0_^6^), then all 10 of these become at large cases, while Kilo through Romeo are exposed but quarantined before they turn infectious. We call Team a cluster of Type 4, and the probability of its occurrence is (1 − *P*_0_*P*_2_) *Q*_0_^4^ (1 − *Q*_0_^6^) *P*_1_. The probability that everyone from Alfa to Juliett is asymptomatic is minuscule and we take it as zero. Thus, we define the Type 5 cluster to be the one where contact tracing has no effect at all i.e. all cases remain at large. The probability of its occurrence is 1 minus all the above probabilities put together. A picture might well be worth the last 500 words, so we present the probability tree below as Fig. 6 and also the case burdens associated with the various cluster types as Table 2. In a similar manner we can account for the probabilities of occurrence of the five types of clusters via SEC events. For this, we replace *P*_2_ by *P*_3_ in the expressions obtained above.

**Figure 6 :**
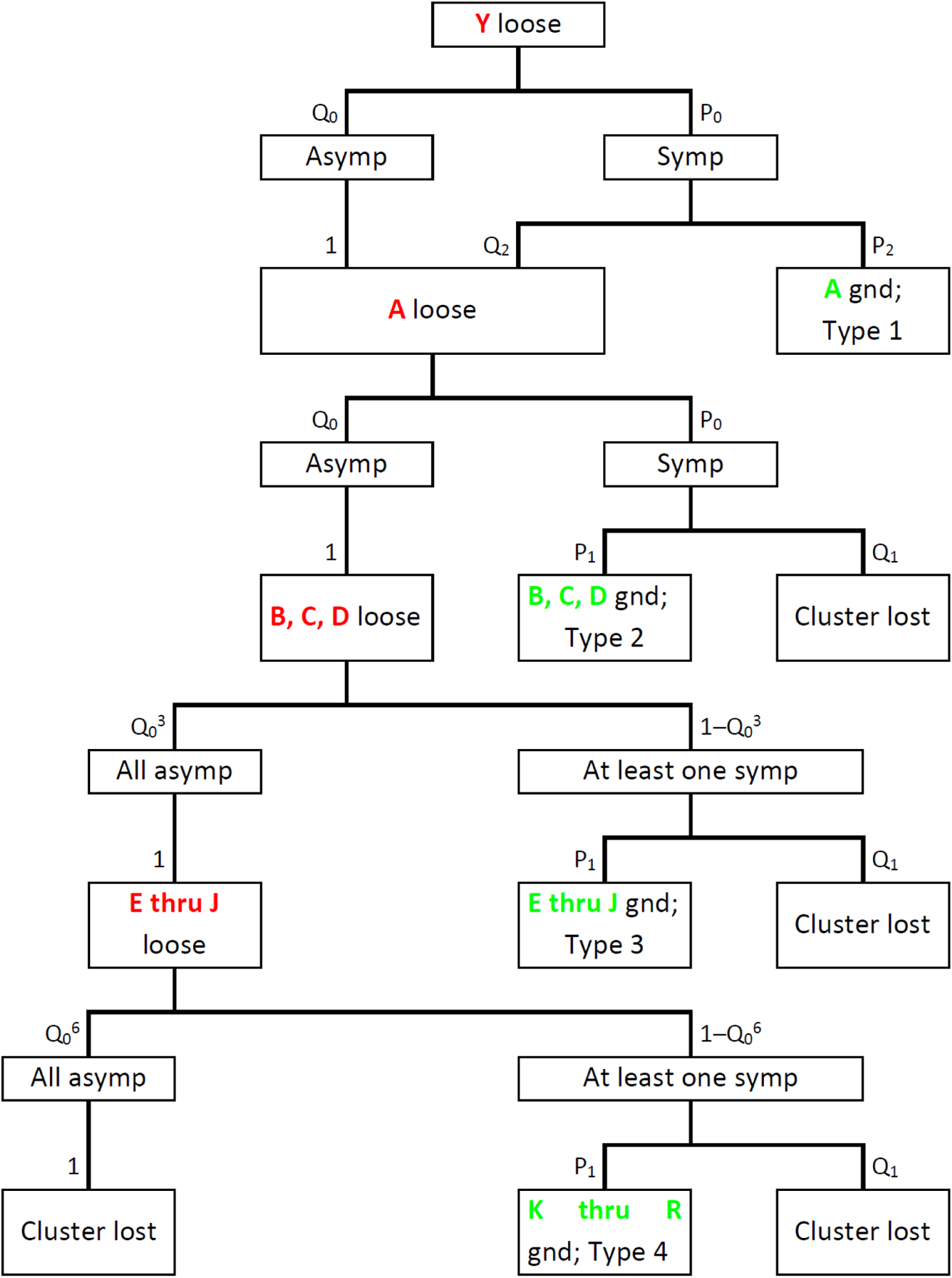
Probability tree showing how Team can end up being a cluster of Types 1 through 5. “Asymp” and “symp” stand for asymptomatic and symptomatic, “loose” means at large and “gnd” or grounded denotes quarantined. “Cluster lost” means that the tracers fail to track down Team i.e. it becomes a cluster of Type 5. The probabilities associated with each event are shown alongside the vertical arrows leading to the event; Q_n_ is shorthand for 1 − P_n_ and certainties are marked as 1 for consistency. If deriving (1) from this diagram, note that Q_0_ + P_0_Q_2_ = 1 − P_0_P_2_.

**Table 2 :** Total numbers of at large and quarantined cases arising from clusters of the five fundamental types.

| Type | 1 | 2 | 3 | 4 | 5 |
| --- | --- | --- | --- | --- | --- |
| At large | 0 | 1 | 4 | 10 | 24 |
| Quarantined | 1 | 3 | 6 | 8 | 0 |

Given these probabilities, we now describe the modifications which must be made to the computational algorithm outlined in Ref. [35]. Firstly, we define extra variables *x_i_* and Δ*x_i_*, the cumulative respectively daily counts of at large cases on day #*i* and *q_i_* and Δ*q_i_*, the same for quarantined cases. The total numbers of socially active cases become *y_i_* = *x_i_* + *q_i_* and Δ*y_i_* = Δ*x_i_* + Δ*q_i_*. The Subroutine roundoff in the algorithm of Ref. [35], which replaces a fraction by its nearest integer and carries over the error, remains as it is. In the main routine, we redefine the total number of at large cases *α* present on day #*i* to be Δ*x_i_*_−1_ + Δ*x_i_*_−2_ + Δ*x_i_*_−3_. The calculation of expectation number *E_U_* of susceptible clusters seeded via UCT on day #*i* remains unchanged from Ref. [35]. This is because contact tracing does not affect the parameters other than *α* which go into determining *E_U_*. Then, we introduce the probabilities obtained above for the seeded clusters belonging to Types 1 through 5. These probabilities, in summary form, are

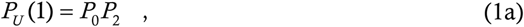

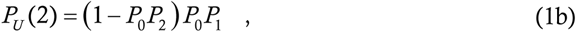

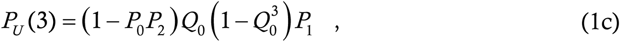

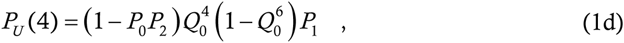

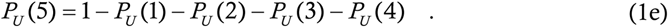

Then, the expectation number of susceptible clusters of type *j* seeded during UCT events becomes *E_U_*(*j*) = *E_U_P_U_*(*j*).

Analogously we calculate the expectation number *E_S_*(*j*) of susceptible clusters of various types seeded on day #*i* during SEC events, by replacing *P*_2_ with *P*_3_ in the expressions above. Adding the two together gives us the expectation numbers *E*(*j*) of the five types of clusters seeded on day #*i*. The sum *E* = *E*(1) + *E*(2) + *E*(3) + *E*(4) + *E*(5) gives the total expectation number of clusters seeded on this day. In Ref. [35] we straightaway rounded this off to get the actual (integer) number of clusters seeded. Here however, there is one more thing to take care of, which is the maximum contact tracing capacity *CT*_max_.

Identification and isolation of a cluster entails tracking down 24 people and issuing quarantine recommendations to all of them. This is time- and resource-consuming work, and needs dedicated personnel. It is reasonable to expect that the city will have an upper limit on the number of contact tracers and hence a ceiling on the number of people who can be tracked down in a day. We call this ceiling *CT*_max_. Now we need an estimate of the number of people required to be traced every day. Generating a cluster of Types 2 through 4 requires identifying 24 people; the count required to generate a cluster of Type 1 is harder to calculate since it involves identifying UCT and SEC transmissions rather than cluster isolation. Nevertheless, for calculational convenience, we will assume that this process involves tracing 24 people also. Generating a Type 5 cluster of course does not require any contact tracing at all. Putting this together, the number of people required to be traced on day #*i* is

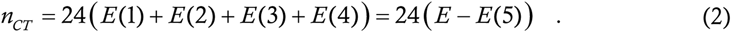

If *n_CT_* ≤ *CT*_max_, then the calculated *E*(*j*)’s for day #*i* are feasible to generate. If this inequality is violated however, then the calculated *E*(*j*)’s are impossible.

In this case, we define the capacity ratio (the early warning indicator) *ρ* = *CT*_max_/*n_CT_* and rescale the expectation numbers of contact traced clusters by *ρ* i.e. we replace *E*(*j*) = *ρE*(*j*) for *j* goes from 1 through 4. This rescaling ensures that the total number of contact tracings made on day #*i* becomes equal to *CT*_max_ instead of exceeding it, while the relative proportions of Types 1 through 4 clusters remain unchanged. To make the rescaling consistent, we define *ρ* = 1 in the case that *n_CT_* ≤ *CT*_max_. The total number *E* of clusters seeded on day #*i* cannot depend on the contact tracers’ ability to find these clusters, since the finding happens only after day #*i*. Hence *E* itself is independent of *ρ* and the adjusted *E*(5) is calculated as *E* less the rescaled *E*(1) through *E*(4). Only now do we perform the roundoff to calculate the integer numbers Δ*z_i_*^(^*^j^*^)^ of susceptible clusters of type *j* seeded on day #*i*.

Another difference from Ref. [35] occurs in the removal of immune clusters. In Ref. [35] we treated entire clusters as susceptible or immune – this was essential to prevent over- or undercounting while ensuring tractability of the mathematical expressions. Here, different types of clusters generate different numbers of susceptible and immune people at the end of the cluster-level outbreak (Type 1 has 23 susceptible and 1 immune, Type 2 has 20 susceptible and 4 immune etc), and treating such clusters as a whole as immune (or susceptible) will lead to unacceptable levels of error. To get around this, we use an approximation scheme where the error involved is much less. We let *w_i_* and Δ*w_i_* (equivalent of *z_i_* and Δ*z_i_* of Ref. [35]) denote the effective numbers of immune clusters. Whenever a Type 5 cluster is generated, we increase Δ*w_i_* by one, just as in Ref. [35]. For the other cluster types, whenever total 24 additional immune people are generated, we increase Δ*w_i_* by one. This scheme ensures that the infection can mathematically spread to the entire population, neither stopping short nor overshooting. Here, we explain why we opted for [1; 3; 6; 8; 5; 1] as the cluster sequence instead of [1; 3; 6; 7; 5; 1] of Ref. [35]. This is because, with this sequence, a Type 5 cluster infects everybody inside and is consistent with our heuristic susceptible cluster calculation scheme, whereas the one remaining susceptible person with the original sequence causes a needless headache at this step.

A final point of difference from Ref. [35] occurs in the treatment of the household cases. In Ref. [35], all socially active cases generate two additional household cases. Here, all at large cases generate the two additional household cases but quarantined cases do not spawn this extra caseload.

### §8. Computer algorithm

We now list the additional variables and parameters present in the model with respect to Ref. [35], and give the algorithm for calculating the case trajectories. In the algorithm, we condense those parts which have already appeared in the algorithm of Ref. [35].

**Table 3 :** New variables and parameters arising when contact tracing is added to the basic CST model.

| Variable | Significance |
| --- | --- |
| $x_i, \Delta x_i$ | Cumulative respectively daily counts of at large cases cropping up on day $\#i$ |
| $q_i, \Delta q_i$ | Cumulative respectively daily counts of quarantined cases cropping up on day $\#i$ |
| $w_i, \Delta w_i$ | Cumulative respectively daily counts of effectively immunized clusters cropping up on day $\#i$. Replaces $z_i$ and $\Delta z_i$ of Ref. [35]. |

| Parameter | Significance | Default value |
| --- | --- | --- |
| $P_0$ | Probability of random case being symptomatic | 0.5 |
| $P_1$ | Probability of grounding cluster upon finding a symptomatic case | 0.8 |
| $P_2$ | Probability of identifying a symptomatic case's secondary UCT transmissions | 0 |
| $P_3$ | Probability of identifying a symptomatic case's secondary SEC transmissions | 0.2 |
| $CT_{\max}$ | The maximum number of people whom the contact tracers can track down every day | 50 |

We have chosen a relatively high default value for *P*_1_ since clusters are composed of close contacts and these are relatively easy to trace. We have gone with zero as the default for *P*_2_ since UCT events like transmission on board buses and inside marketplaces are almost impossible to identify in practice. For *P*_3_ we have gone with a low default value since SEC transmissions, for example at wedding and birthday parties, are harder to identify than intra-cluster transmissions but easier than transmissions in random public places.

We now give the algorithm itself. The algorithm is schematic and is not cast in any particular language. Our code is written in Matlab and is being included in SI.

**Algorithm 1 :**
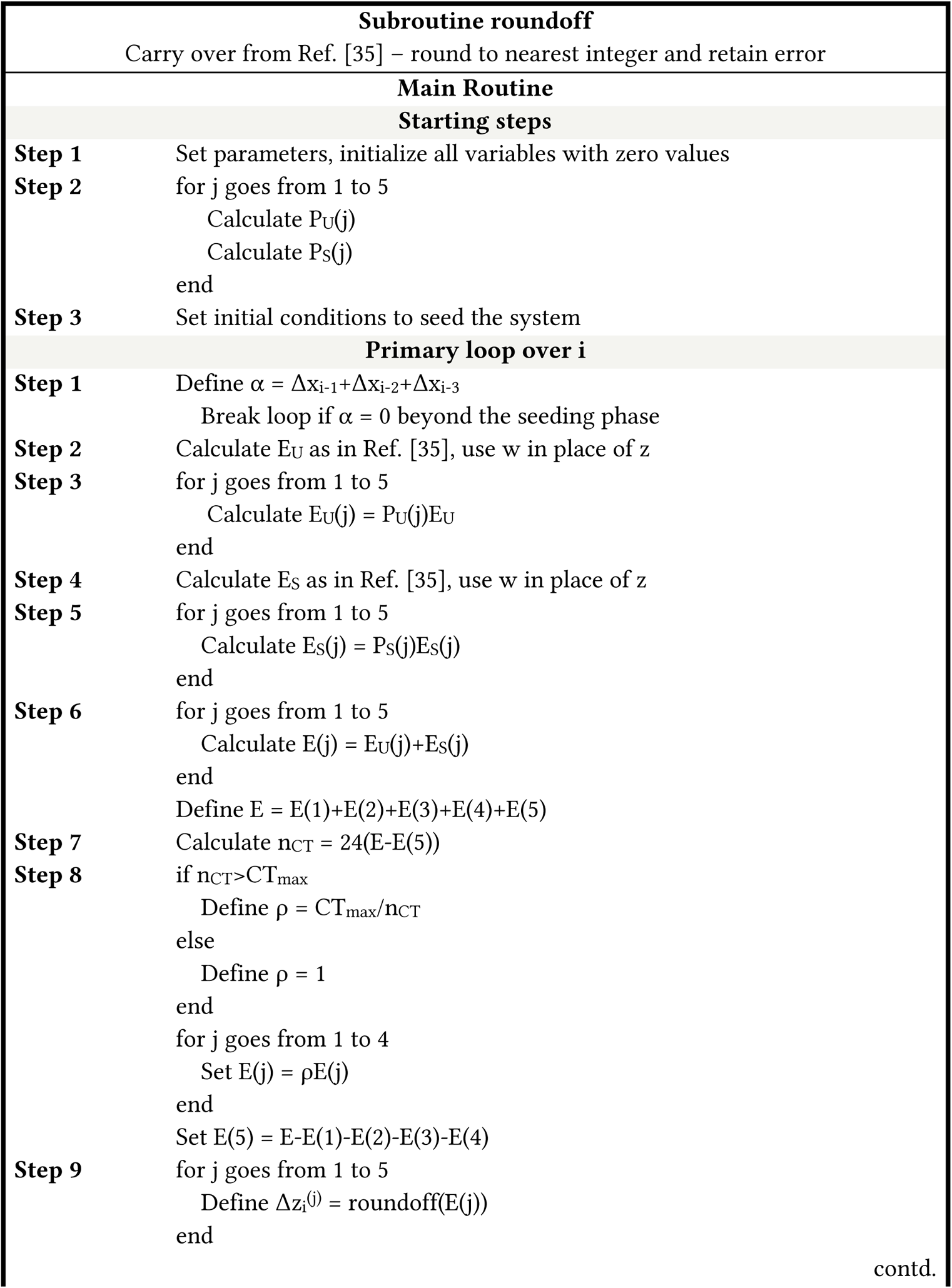
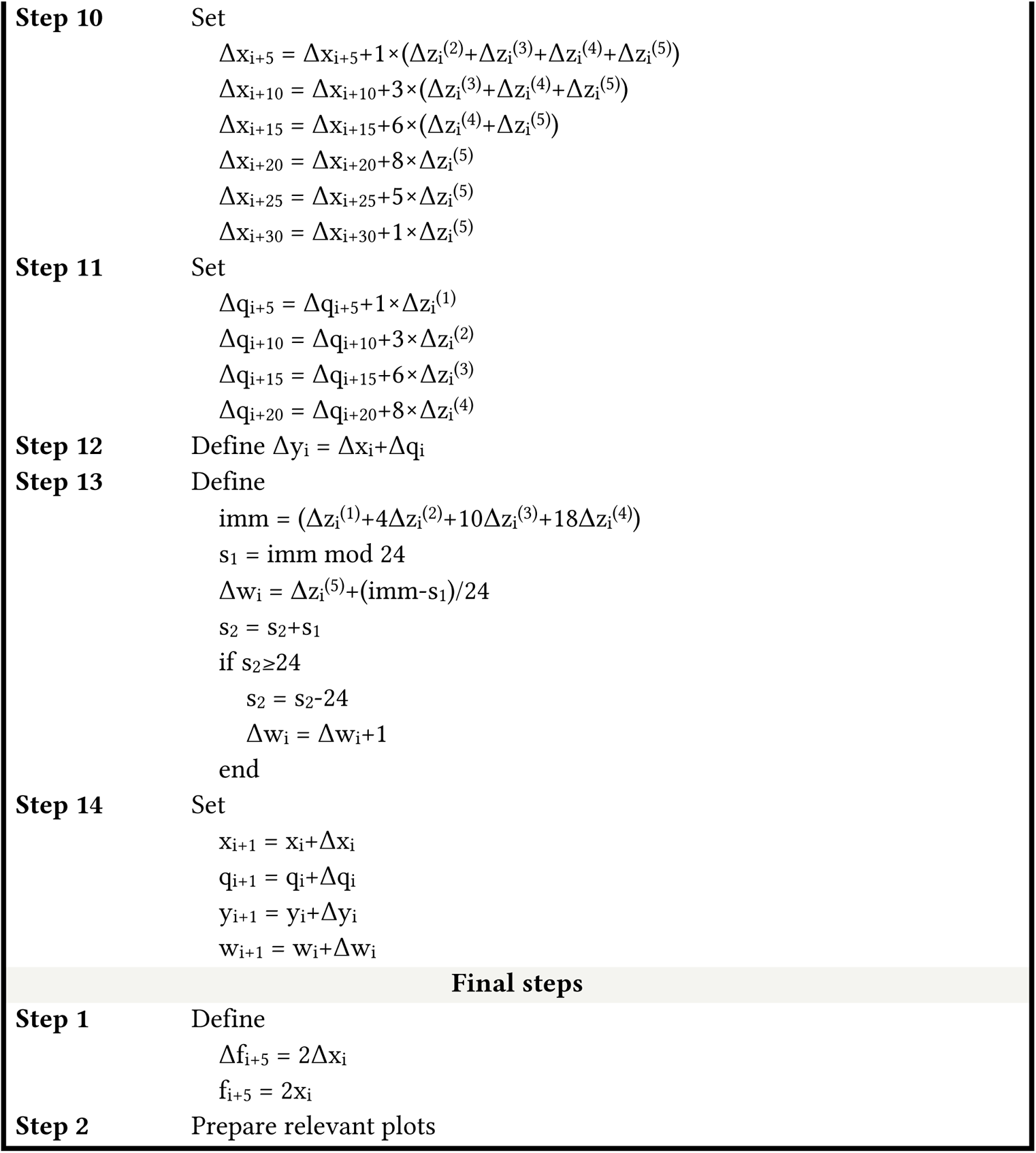
Schematic form of the routine used to compute case trajectories. imm in Step 13 of the primary loop counts immune population. Steps 10, 11 and 13 make explicit reference to the sequence [1; 3; 6; 8; 5; 1] but that is only for readability – it is trivial to replace them by v(1), v(2) etc where v denotes the cluster vector. Note that the numbers 1, 4, 10 and 18 in Step 13 are v(1), v(1)+v(2), etc.

As in Ref. [35], we introduce the parameter *k*_max_ for convenience and set its value to 80. In all simulation results, we use the initial condition that eight clusters are seeded on the first day, after which the disease evolves on its own.

### §9. Limitations of the model and their consequences

Most of the limitations of the model have already been mentioned in Ref. [35]. They are

- Assumption of constant cluster size
- Assumption of constant cluster sequence
- Use of roundoff
- Decoupling of households from clusters
- Ignoring the possibility of one person’s belonging to multiple clusters

Reference [35] also discusses the ways of circumventing these assumptions – in short, we have to convert the deterministic model to a fully agent-based model. The additional part implementing contact tracing contains two assumptions which we have already discussed in §7. There is an approximation in the calculation of *ρ* since we assume that all the cluster members are being traced on the same day as the case is found. In reality, the most immediate contacts might be identified first and the more distant ones later, easing the tracers’ job. On the average however, the total number of contacts needing to be traced over say a week or ten days will remain unchanged. Another approximation arises in our counting of immune clusters *w_i_* and Δ*w_i_*. In this case, since our scheme preserves the total susceptible and immune populations at any time, errors will tend to average out.

Once again, converting from the deterministic CST to a fully agent-based model will obviate the need for most or all of these assumptions. A more interesting question is : how might our assumptions influence the model resultsAs a first test, we have varied the parameter which might be the most restrictive, which is the cluster sequence. Instead of [1; 3; 6; 8; 5; 1], we have now considered the sequence [1; 4; 13; 6]. This describes much faster intra-cluster transmission, corresponding to a more transmissible mutant strain. Concomitantly we have also increased the values of *P_U_* from 0·15 to 0·25 and *m_S_* from 2 to 3·5. Note that these values are hypothetical and we neither state nor intend to suggest that the percentage increases in transmissibility parameters are actually representative of B1.617.2 or any other actual SARS-CoV-2 variant. In this case, we have run extensive simulations and found all the effects mentioned in §2. The parameter values at which the different behaviours occur are of course different, but that apart everything else is the same. We dispense with another plethora of figures, since the code is already available in full in SI.

A realistic society is expected to have a distribution of cluster sizes and sequences as well as transmission parameters, and not just constant values of all of them. At one end of the spectrum, college students for example will tend to have large clusters, high intra-cluster interaction resulting in a rapid sequence, as well as higher values of *P_U_* and *m_S_* arising from their active social lives (and possibly lack of caution). At the other end, retirees are expected to have small clusters, a less sharp cluster sequence and low *P_U_* and *m_S_* due to their relatively limited social lives as well as justified fears about the consequences of contracting the disease. In such a society, we expect some of the special effects to remain more or less as is and others to be altered.

The critical mass effect ought to be robust because of its mechanism of action. With parameters for most or all sub-populations in a dangerous region, when the college students are operating at the borderline of tracing capacity, cases in that sub-population will plateau; cases in less contagious sub-populations will also plateau or decay (recall that in CST model, the plateau is manifest in large regions of the parameter space). The students will be the first to cross the contact tracing threshold and activate the instability; cases arising there will then induce cases in the other sub-populations and cause them to follow suit as well. On the other hand, a distribution of cluster size and contagiousness will certainly blunt the edge of the knife. When an NPI is progressively relaxed, the critical parameter value for the college students will be lower than that for the retirees, and crossing that first cutoff alone will not initiate a wave in the whole population. For a mass transition from plateau to wave, we shall have to effect a larger change in parameters than Fig. 2 would imply. Like critical mass effect, hysteresis effect is again expected to be robust in a population with a distribution of cluster sizes and parameters, since it will apply individually to the various sub-populations. As we have mentioned in §3, demonstrations of the critical mass and hysteresis effects have indeed occurred many times in different parts of the globe. Finally, we mention that the consequences of errors arising from approximations in our implementation of the model (for instance, use of the parameter *k*_max_) will be no different from those of approximations in the model itself. We believe that the undulation in caseload in Fig. 2 as *n_S_* is increased beyond the knife-edge is an example of this kind of error – without *k*_max_ or roundoff for instance, the curve here would likely have been monotonic, which is more realistic.

A further limitation exists with respect to data fits. Here we have not presented any fits or predictions of COVID-19 curves for actual regions. This is because the model has a lot of free parameters whose real-world values we do not know, key among them being the cluster size and sequence, *n_U_*, *P_U_*, *n_S_* and *m_S_*. Without further information, good fits to data shall become generated in many regions of the parameter space. To make the fit meaningful, we shall have to obtain estimates of as many of the parameters as possible from sources external to the COVID-19 trajectories themselves. In Ref. [35] we have argued that, since the bulk of the parameters relate to people’s interaction, the contact tracers’ logs provide an ideal source from which to estimate their values. Unfortunately, we currently lack access to such logs and are not aware of even one repository where they are publicly available. Hence, for the timebeing we stick to universal, parameter-independent results and leave region-specific fits and predictions for a future study.

## Data Availability

This Article does not use data. All simulation code is available on demand.

## Supporting Information

### §S1. Demonstrations of the knife-edge effect

In Fig. 2 of the Article proper we presented the knife-edge effect for variation of *n_S_*, with all other parameters being held to their default values as mentioned in §7-8. Here we show the same thing for variation of *P*_1_ and *CT*_max_. As in Fig. 2, the epidemic final size is a blue line attaching to the left hand *y*-axis while the duration is a green line attaching to the right hand *y*-axis. Recall that the total population of the city is 3,02,400.

**Figure S1 :**
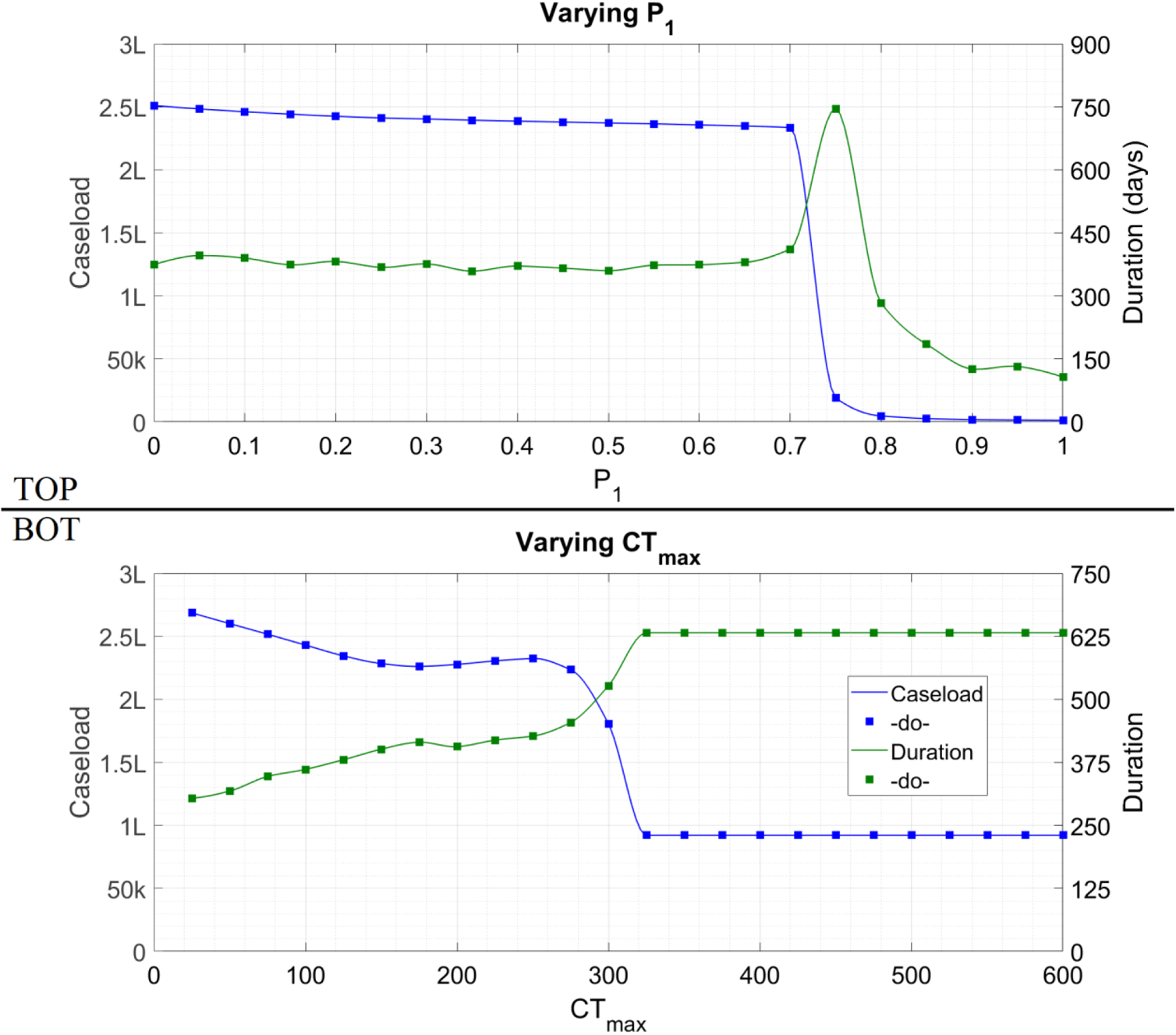
Cumulative caseload (blue) and duration (green) of the epidemic as the labelled parameter is varied and all other parameters held to their default values. The symbol ‘k’ denotes thousand and ‘L’ hundred thousand.

In §9 of the Article proper we have mentioned the undulations in Fig. 2 as the likely result of computational inaccuracy. The undulations in case count in Fig. S1-bot to the left of *CT*_max_ = 320 are very likely a similar phenomenon. The perfectly flat line to the right of this *CT*_max_ is not an error however since for all *CT*_max_ above this value, every contact who can be traced is traced and the epidemic evolution remains unchanged.

### §S2. Demonstrations of the hysteresis effect

We include here the simulation plots for the situations mentioned in §2 of the Article proper but not explicitly shown. For the first situation we start from *n_S_* = 1900 and implement stronger NPI by reducing *n_S_* on day #100. Noting from Fig. 2 that the knife-edge lies between 1750 and 1800, we consider a mild reduction to 1500, as well as a much stronger reduction to 300. As in Fig. 3 of the

Article proper, the daily new cases are grey bars attaching to the left hand *y*-axis while the cumulative cases are a blue line attaching to the right hand *y*-axis.

**Figure S2 :**
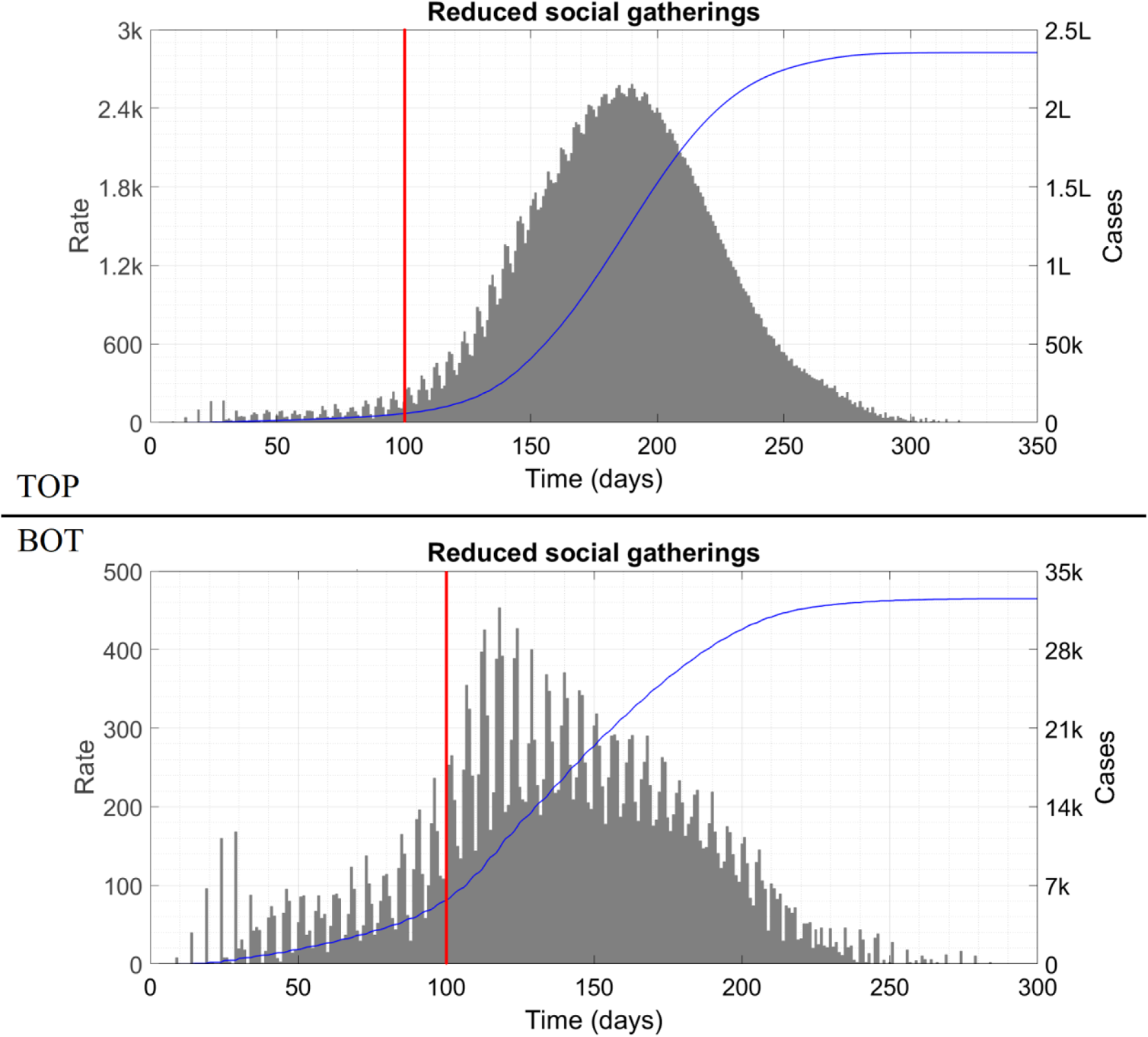
Top panel shows the effects of reducing n_S_ from 1900 to 1500 while bottom panel shows the effects of reducing it to 300. In both cases, the intervention is applied at 100 days, indicated by the red vertical line. The symbol ‘k’ denotes thousand and ‘L’ hundred thousand.

Even though *n_S_* = 1500 is acceptable if implemented from the get-go, it fails miserably when applied partway into the wave. This is qualitatively similar to what happened in Maharashtra, as discussed in the Article proper.

In the second situation we initiate the wave with *n_S_* = 1900 and implement an increase in *CT*_max_ from 50 to 400 at 120 respectively 100 days.

**Figure S3 :**
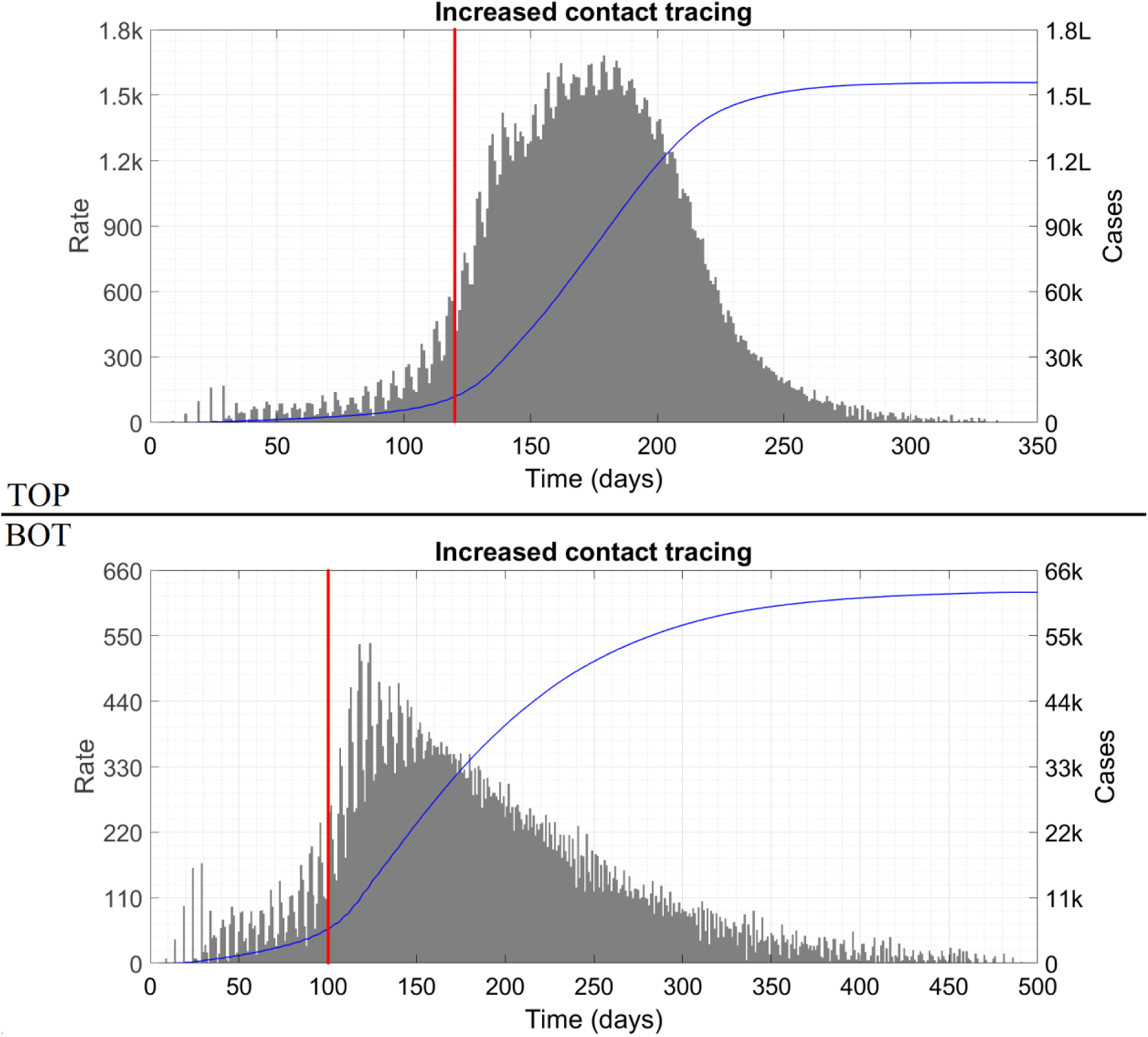
Effect of increasing CT_max_ from 50 to 400 at day #120 (top panel) and day #100 (bottom panel). In both cases the intervention is depicted as a red vertical line. The symbol ‘k’ denotes thousand and ‘L’ hundred thousand.

These plots show that, like the knife-edge effect, hysteresis effect also occurs for variation of different parameters.

### §S3. Discussion of *ρ* as an early warning indicator

In this Section we present plots showing the correlation between a drop in *ρ* and an imminent wave for a wide variety of situations. In Fig. 4 of the Article proper we have shown this for two contrasting situations taken from either side of a knife-edge. Here we consider the situations which demonstrated the hysteresis effect, and analyse whether *ρ* can tell us in advance if the intervention/relaxation has been well-timed or mistimed. First, we take Fig. 3 of the Article proper, zooming in to focus on the most relevant part (near the unlock). In this Figure and the ones that follow, we show *ρ* as a blue line attaching to the right hand *y*-axis.

**Figure S4 :**
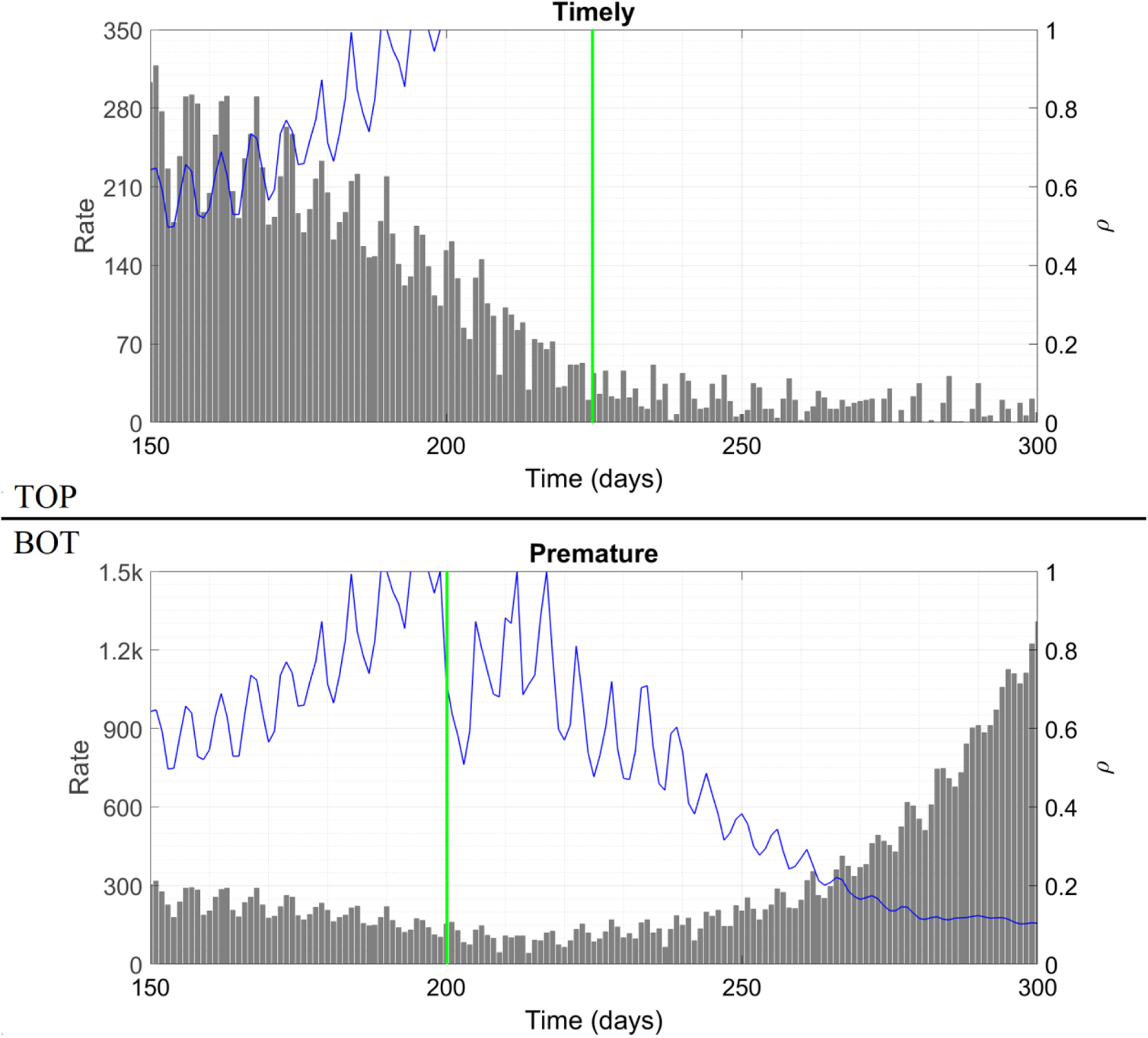
Case trajectories and capacity ratio as function of time for an increase in n_S_ from 300 to 1500 being applied at day #225 (top panel) and day #200 (bottom panel). The point of relaxation of the intervention is shown as a green vertical line.

Next we consider the two situations of Fig. S2.

**Figure S5 :**
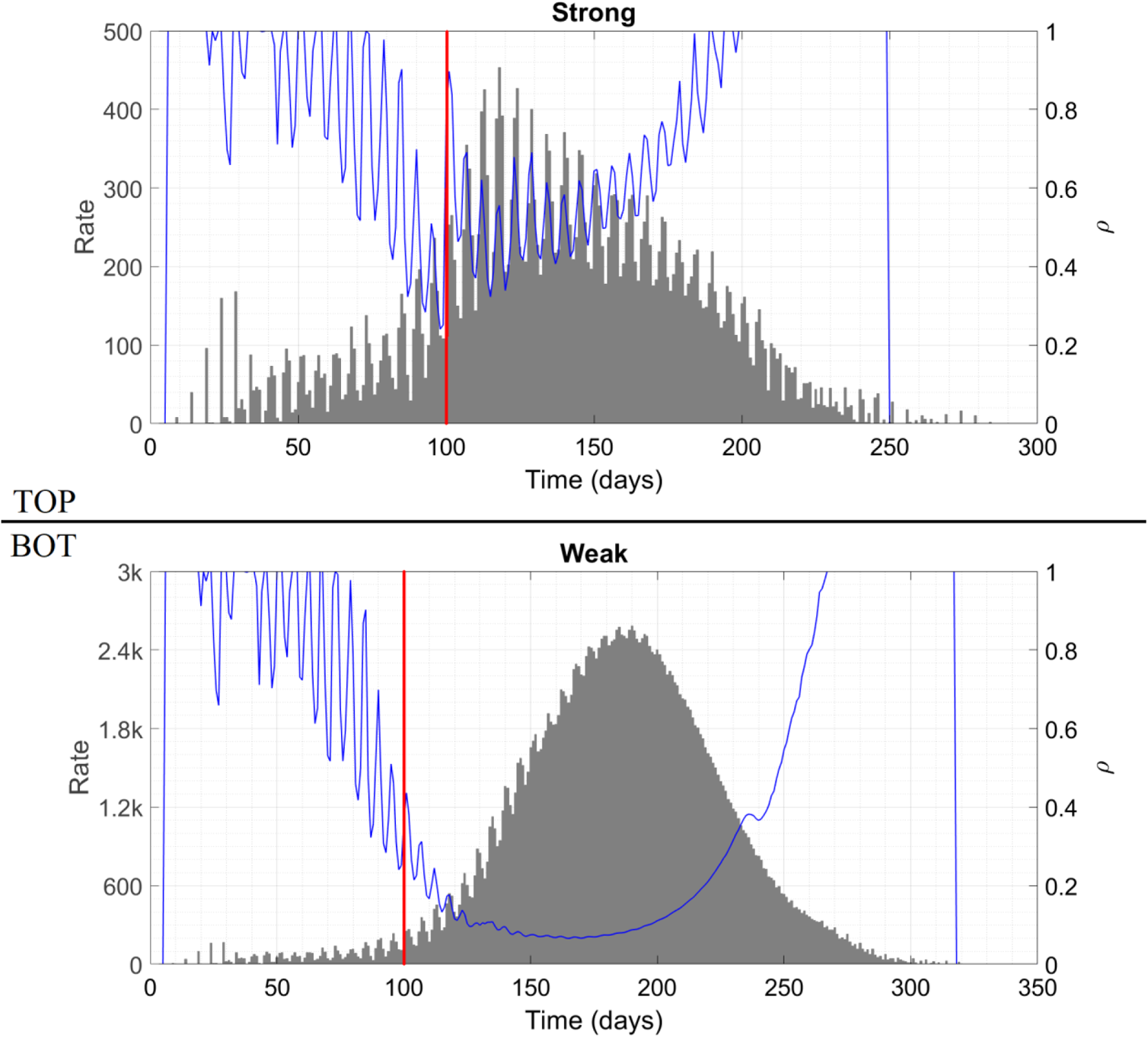
Case trajectories and capacity ratio as function of time for a reduction in n_S_ from 1900 to 1500 (top panel) and 300 (bottom panel) being applied at day #100. The point of application of the intervention is shown as a red vertical line. The symbol ‘k’ denotes thousand.

Next, the two situations of Fig. S3.

**Figure S6 :**
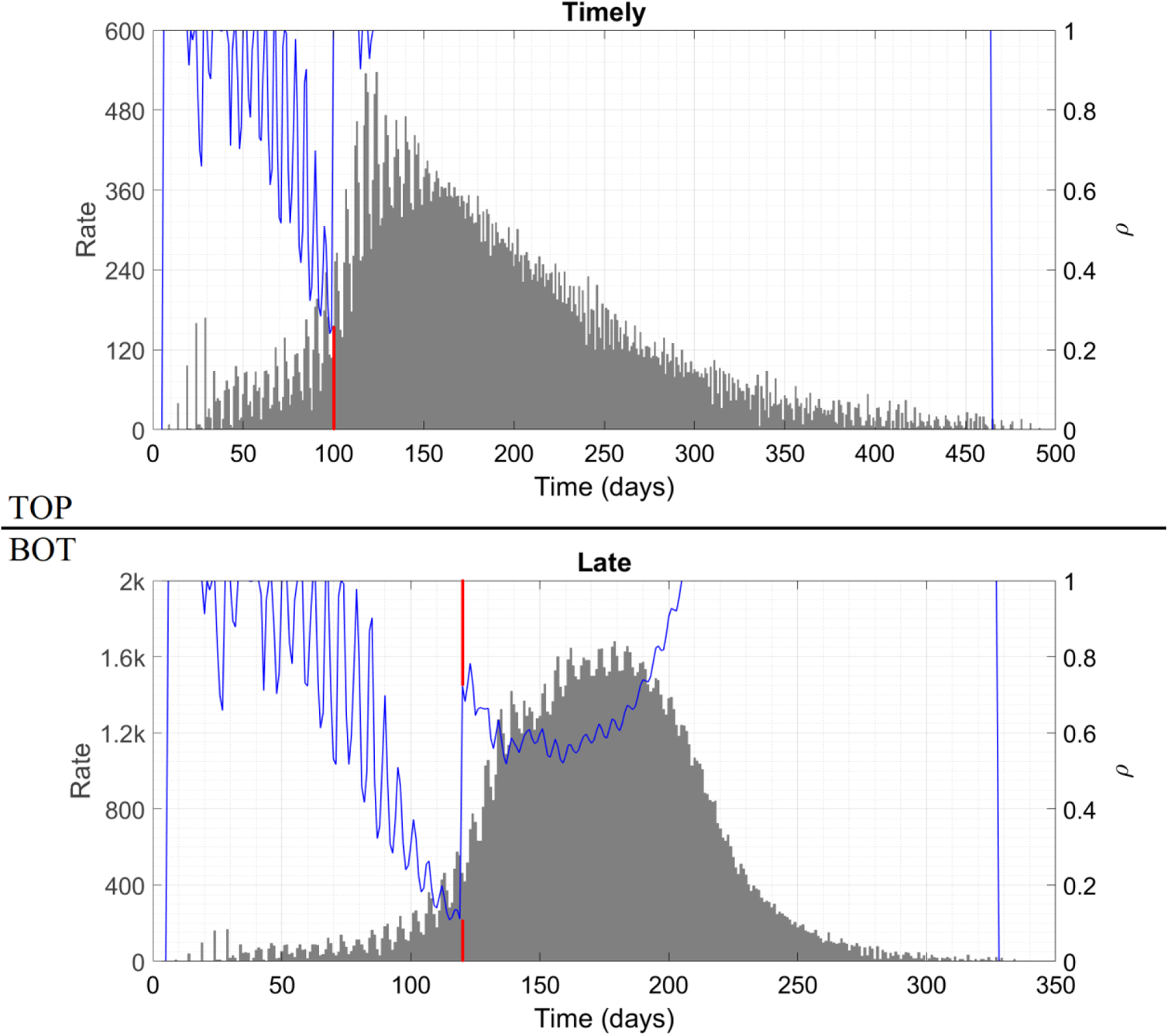
Case trajectories and capacity ratio as function of time for an increase in CT_max_ from 50 to 400 being applied at day #100 (top panel) and day #120 (bottom panel). The point of application of the intervention is shown as a red vertical line. This line is broken because it interferes with the curve of ρ which has a jump occurring at the same day. The symbol ‘k’ denotes thousand.

As our final example, a rerun of Fig. 4 of the Article proper but with the more transmissible mutant mentioned in §9. We also set *CT*_max_ to the value 100. This time, the knife-edge lies between *n_S_* = 750 and *n_S_* = 800. Below are the initial time traces for these two situations.

**Figure S7 :**
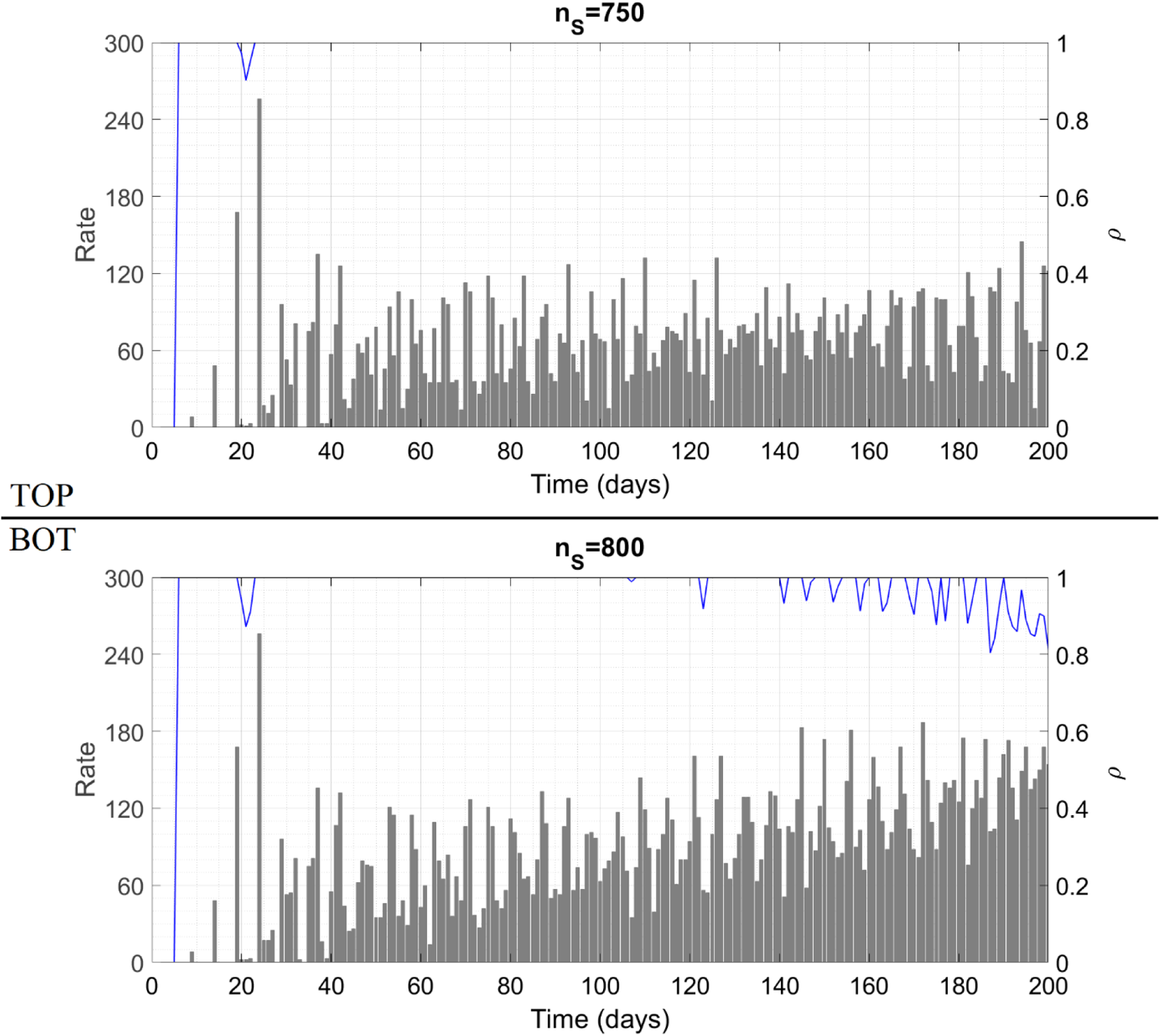
The initial portions of case trajectories and capacity ratio as a function of time for two contrasting situations.

This is almost identical to what we saw in Fig. 4. In the subsequent evolution not shown here, the top panel proceeds almost unchanged to a cumulative caseload of 32,000 while the bottom panel takes off at around day #230 to reach a maximum daily rate of 2500 cases and a cumulative caseload of almost 2,30,000.

We can now give our observations. In all of the five Figures (4, S4-S7), the unfavourable case features a plateau with *ρ* < 1 transitioning into a wave, or an existing wave with *ρ* < 1 accelerating further to mammoth proportions. Thus we can conclude that *ρ* < 1 is an indicator of imminent danger. In four of the five situations (exception is Fig. S5), in the favourable case *ρ* increases to or remains at unity during the critical phase and the epidemic proceeds to a happy conclusion. In Fig. S5-top, *ρ* does increase significantly after the lockdown but fails to attain the maximal value; even so, the wave is crushed by the severity of the lockdown. Thus, if we use *ρ* as a trend indicator, we might get a false alarm in this instance. Note however that from a public health perspective, false alarms are merely a nuisance while the converse – missing an impending escalation – can have tragic consequences. Extrapolating from Fig. S5, we might be tempted to calculate or assign a “safe lower bound” for *ρ* which is less than unity. This however is dangerous as Fig. S7 shows – in this case, the unfavourable situation features a much higher *ρ* than Fig. S5-top. This smaller departure from unity is sufficient to set off the wave in Fig. S7-bot because (*a*) the viral strain here is more transmissible, and (*b*) *CT*_max_ here is higher so a smaller deviation corresponds to a larger absolute count of at large cases. For this reason, the only acceptable value of *ρ* is strictly unity; this is not too stringent a condition since *ρ* cannot exceed 1 by definition.

Thus, extensive simulation results bolster our claim of efficacy of *ρ* as an early warning indicator. Even though many parameters – *n_U_*, *P_U_*, *n_S_* and *m_S_* to name just a few – influence the epidemic trajectories in subtle and diverse ways, the bottom line is that whether a wave will begin next week depends entirely on the capacity ratio this week. In our analysis we have defined *n_CT_* (a constituent of *ρ*) in terms of the model social structure and assumptions. This parameter can however be calculated and/or measured for any social structure. By now, the contact tracing authorities of most regions have a fair idea of how many contacts they need to track down per case to keep the disease in check – for example, an Australian study [42] reports that 25,300 contacts were traced for 619 cases during a period of decline and plateau, translating to average 40·9 contacts per case. This average multiplied by the daily case count approximately gives *n_CT_*. Deciding whether *ρ* is at or below unity in a real world scenario almost does not require any calculation – the moment the contact tracers feel that due to time or logistical constraints they are being unable to thoroughly investigate every emerging case, *ρ* has decreased below unity.

We expect that *ρ* will also be related to the test positivity rate (TPR) in some way – lower *ρ* is expected to correspond to higher TPR. However, the exact nature of the association will depend significantly on the region’s testing policy for identified contacts, as well as walk-in symptomatic persons. Relying on TPR rather than *ρ* will also result in pointless delay of one serial interval before taking action, since the positive tests are expected to come out one serial interval after the exposures are identified. Hence we stick to *ρ* rather than TPR as the early warning indicator.

### §S4. Cautionary points for an efficient contact tracing system

In the Article proper, we have suggested an extensive contact tracing program as a key measure in combating the spread of COVID-19. To increase the efficiency of the program, the authorities might need to keep two things in mind.

Firstly, a large-scale contact tracing program might have an unpleasant corollary in that many people may be treated as potential cases and quarantined despite not actually having been exposed to the virus. In our example of §7, if Team is a Type 1 cluster then only Bravo, Charlie and Delta are actual cases while Echo through Xray are also grounded out of abundance of caution. Thus, in this instance, there are more than six times as many quarantine orders issued as are necessary. For high-interaction persons like shopkeepers, such false alarms might be raised every few days. This will cause economic harm, undue mental stress as well as public lack of faith in the contact tracing system. To mitigate this, two options are available. The first is for asymptomatic suspected exposures to perform a daily self-test, the logistics of which as a quarantine substitute have been discussed in Ref. [41]. The second is to not ground the person entirely unless symptomatic but to ensure that over the next few days s/he follows 100 percent adherence to COVID-19 protocols at work and essential activities, and refrains from inessential activities. Data from Cornell University’s extensive case surveillance network has not found even one instance of in-classroom viral transmission, suggesting that a ‘soft’ quarantine may be as effective at preventing transmission as a ‘hard’ one.

The second point to remember is that many people might be unwilling to confess mask or separation violations to the contact tracers, for fear of legal repercussion and/or adverse judgement. Such false testimony will reduce the efficacy of the tracing program, and foster the spread of the disease. To elicit truthful information, there will have to be well-publicized amnesty policies for instances of COVID-inappropriate behaviour determined from a contact tracing investigation (as against from say enforcement patrols). The tracers will also have to possess communication skills to not sound judgemental, critical or moralistic while asking or receiving answers to questions concerning people’s behaviour.

### §S5. Matlab codes used to generate the results

We give here the Matlab code which generates Fig. 1 of the Article proper. The codes for the remaining figures are trivial variations on this program. Since we have already given the algorithm in detail, the code itself might appear somewhat terse in places. The code is in a form that if you follow the accompanying instructions then you will be able to generate Fig. 1 at the end.

First, please copy the following, paste it into a Matlab window and save it in a folder of your choice as the function file `prob1.m`. Although the code here reproduces Matlab’s default indentation, indentation may be lost when copying from the pdf. In this case, please use the smart indent feature (Ctrl+I) to improve the indentation. This instruction remains valid for all the pieces of code given here.

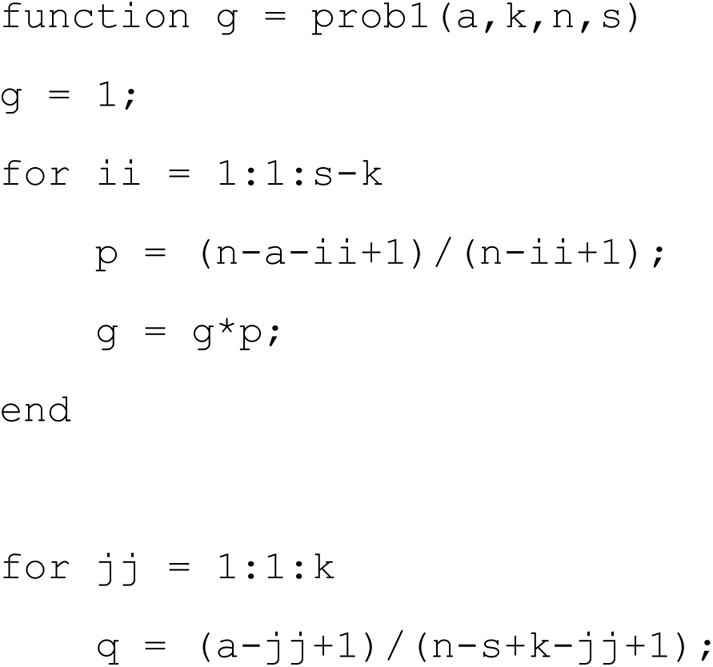

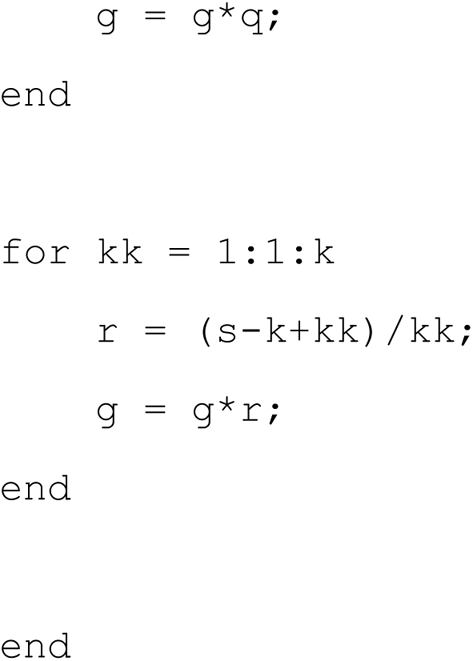

Next, copy the following and save in the same folder as the function file `prob2.m`.

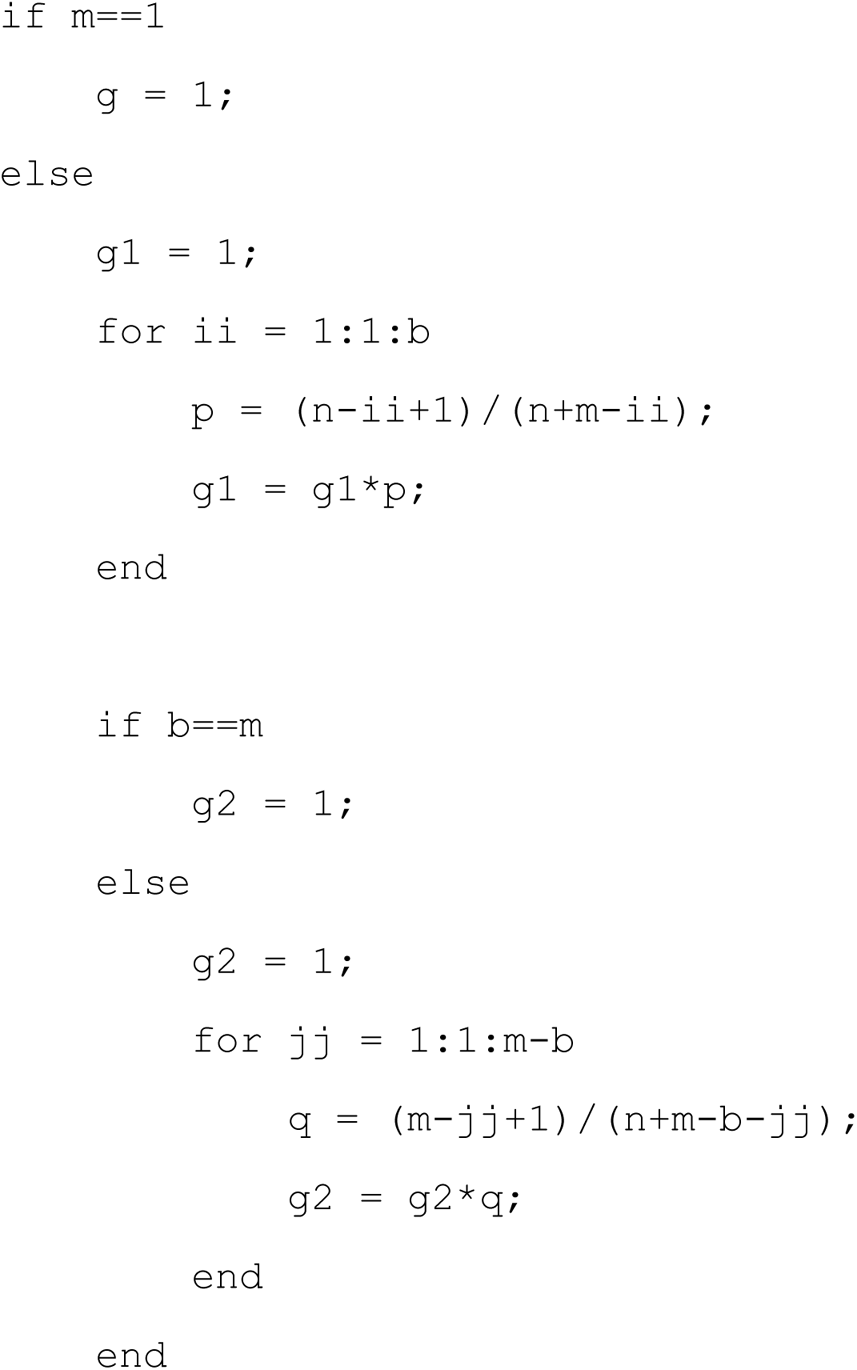

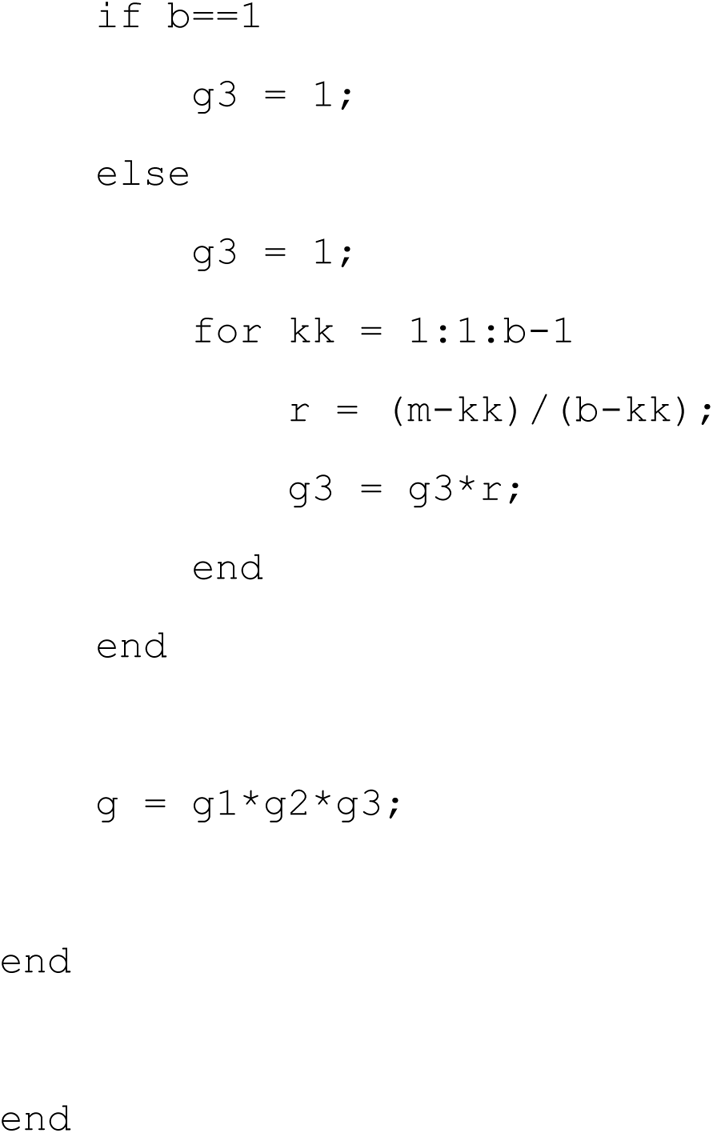

Next, please do the same with the function `prob3.m`, below.

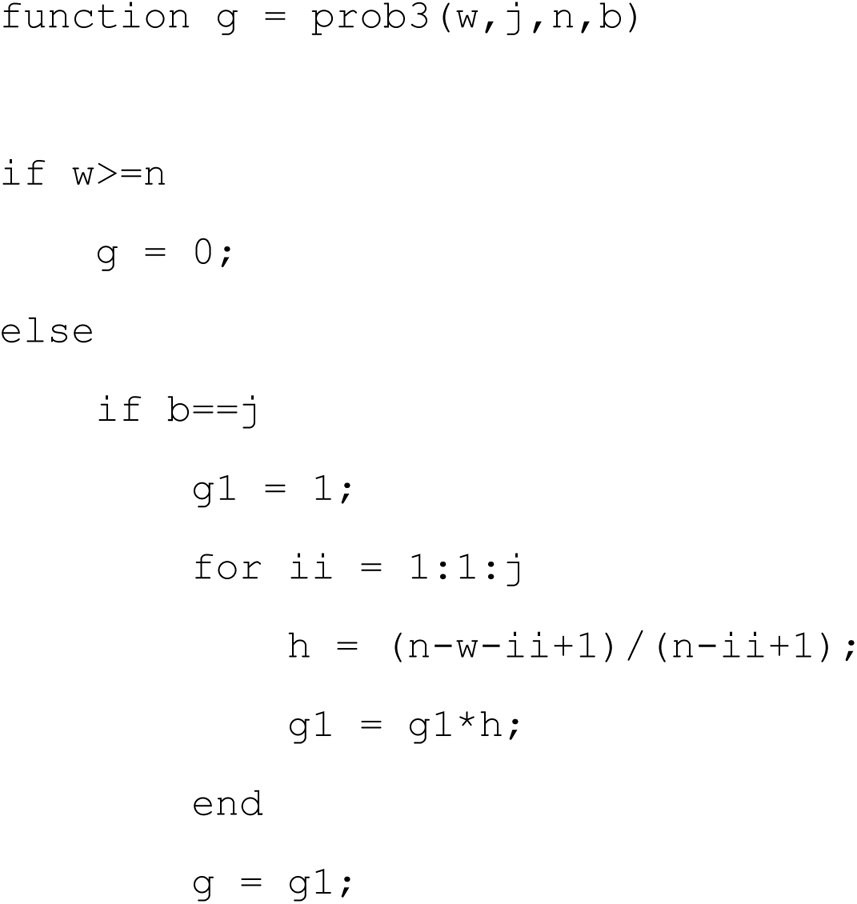

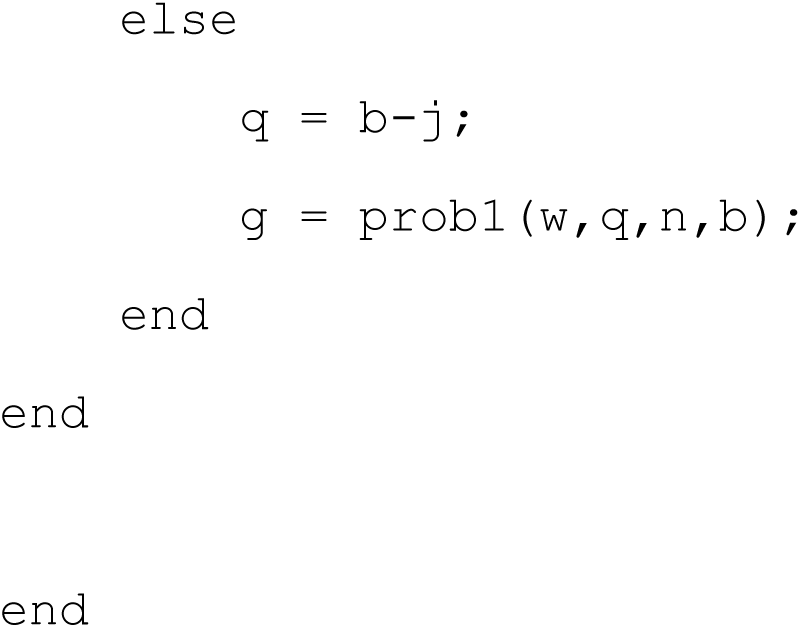

The purpose of these three functions is to make feasible the evaluations of the probabilities involved, as has been discussed in the Appendix of Ref. [35]. After this, please copy and paste the following code and save as the function `roundoff.m`.

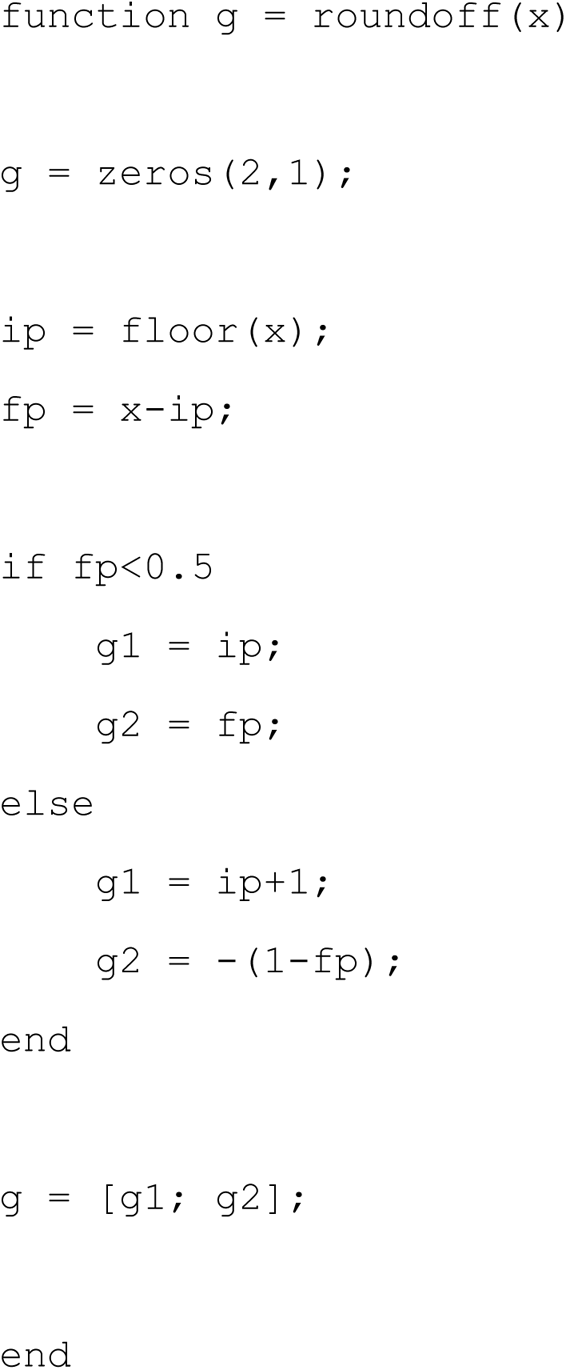

With these preliminaries completed, please copy and paste the following routine into the matlab window and save it in the same folder as a script with any filename of your choice. This routine implements Algorithm 1.

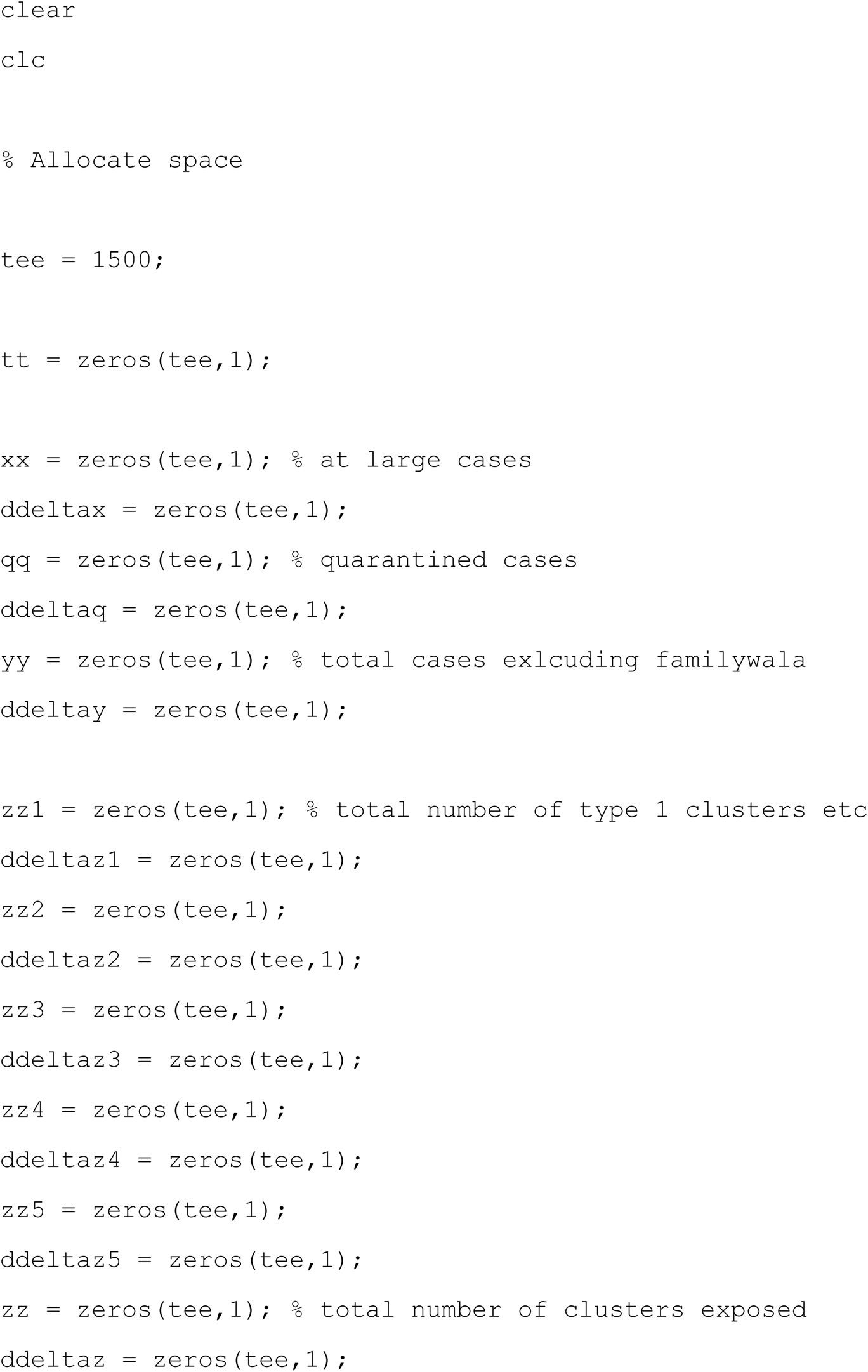

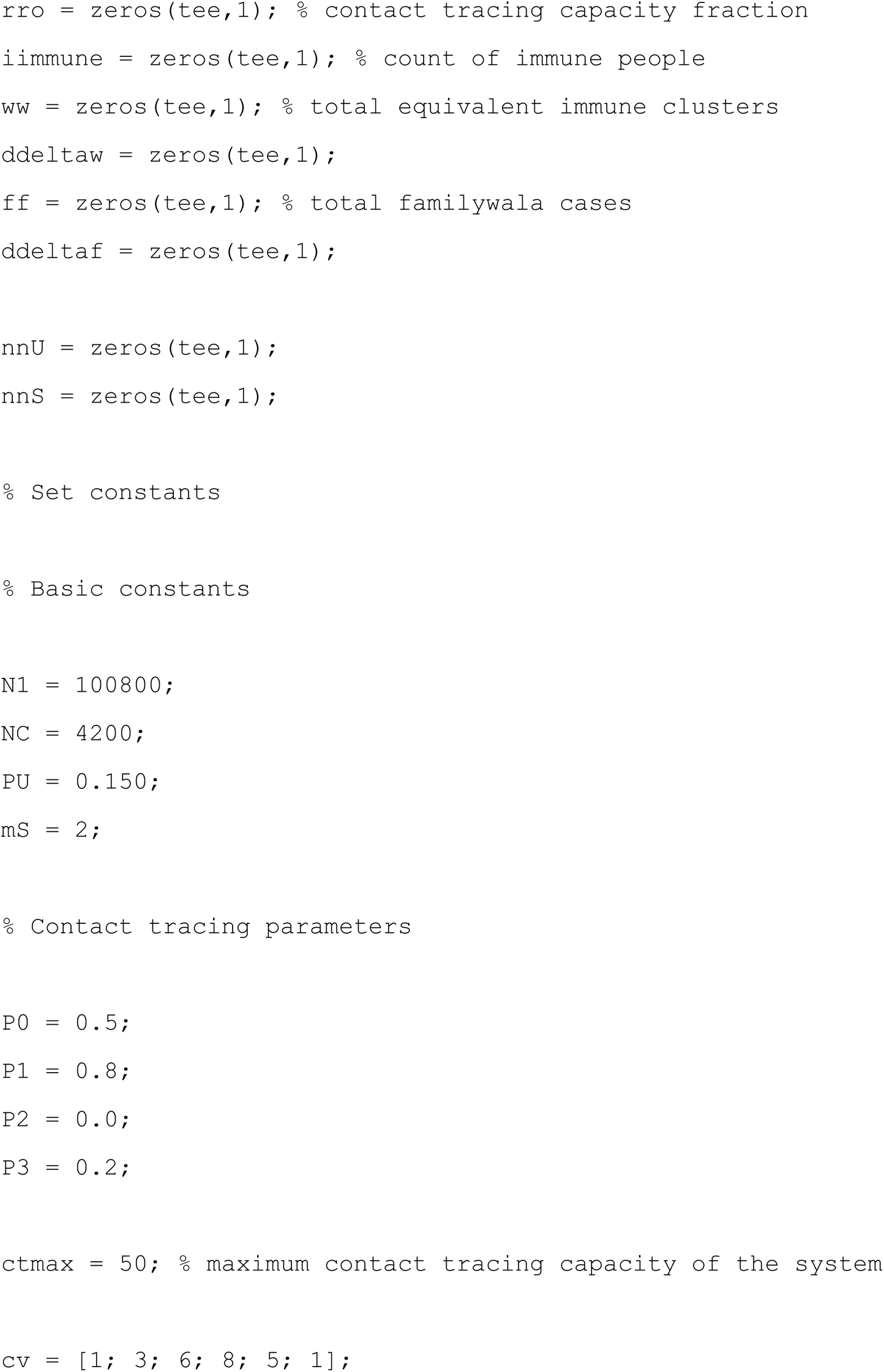

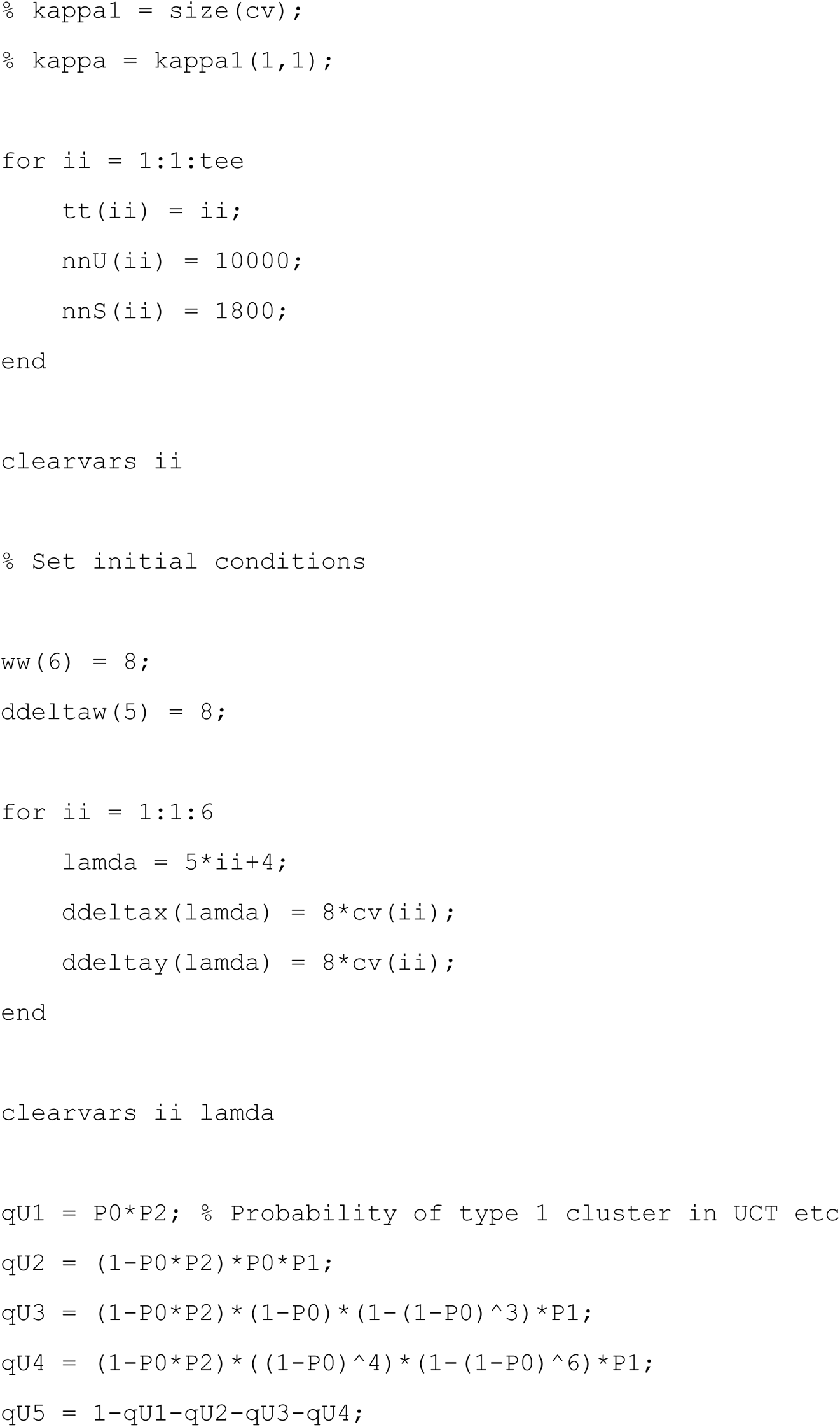

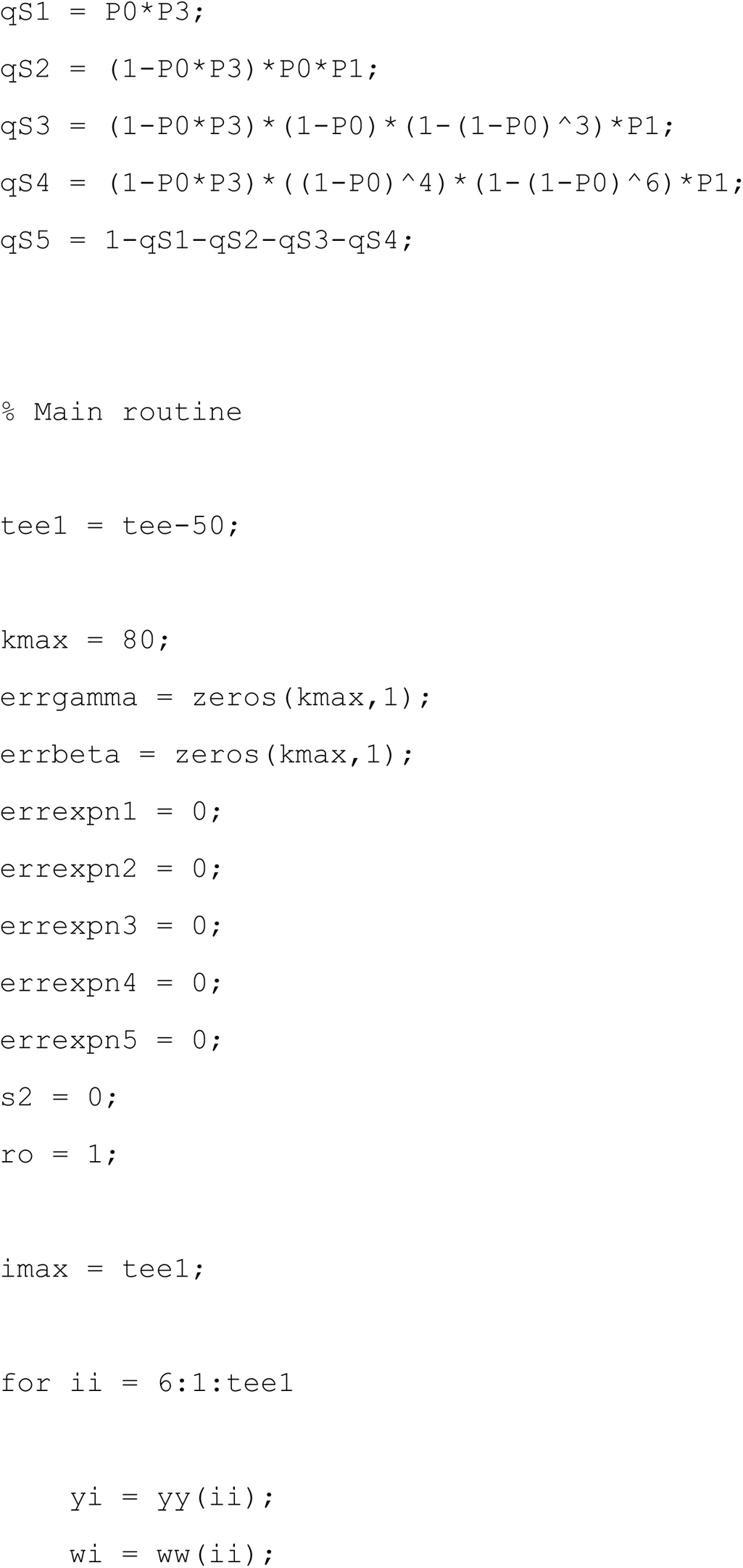

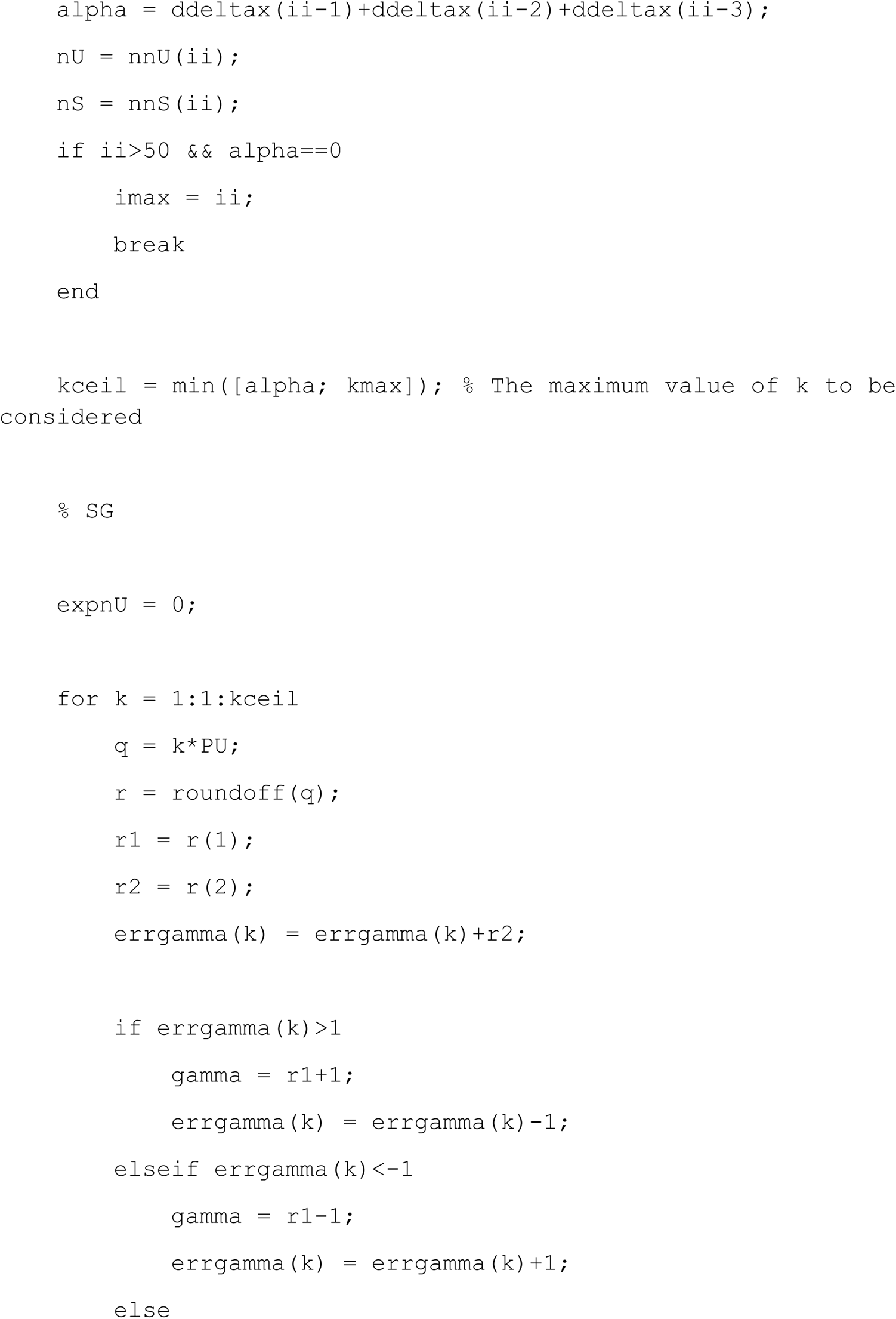

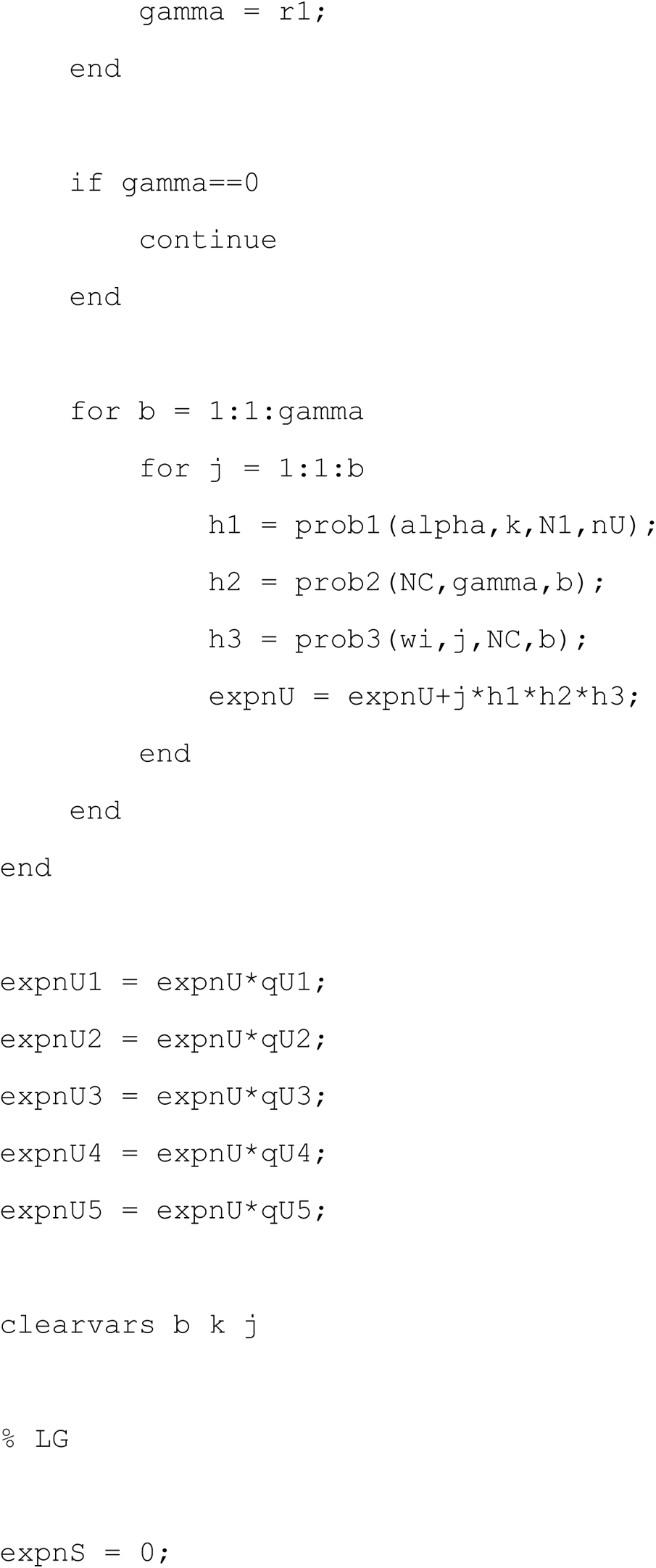

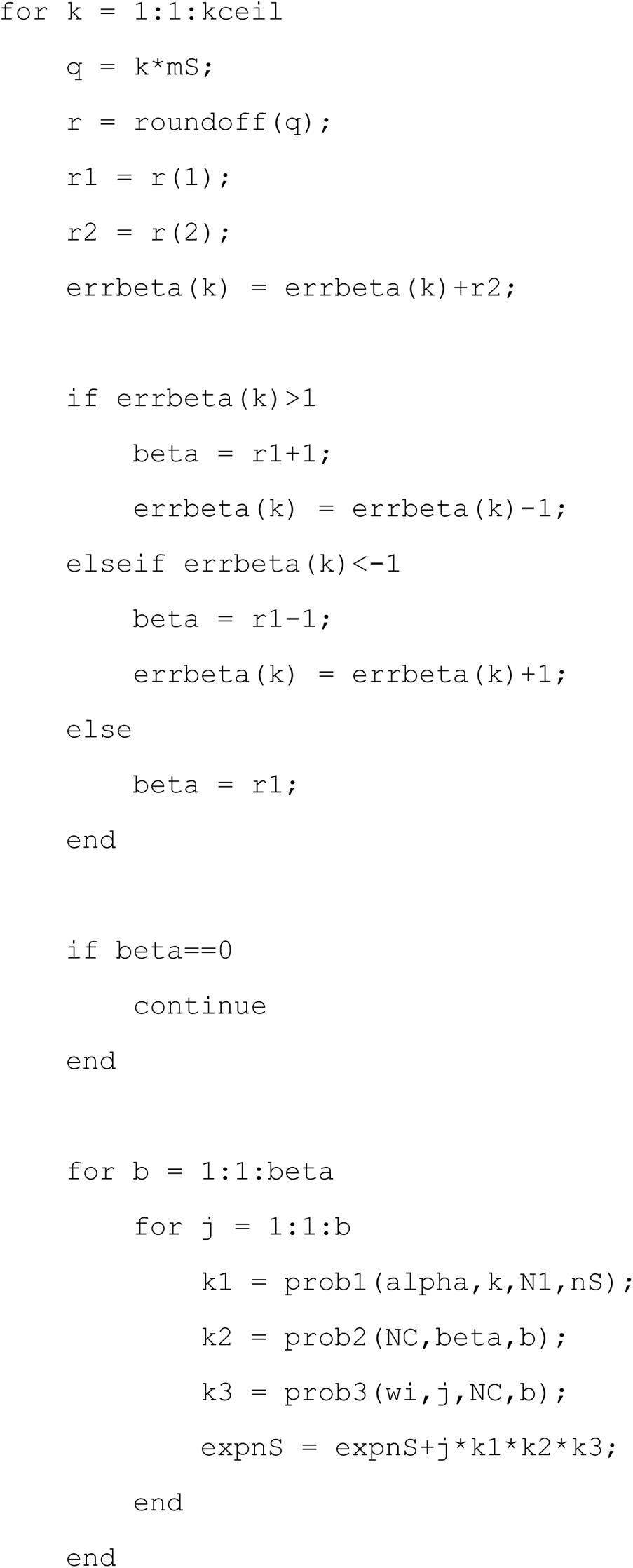

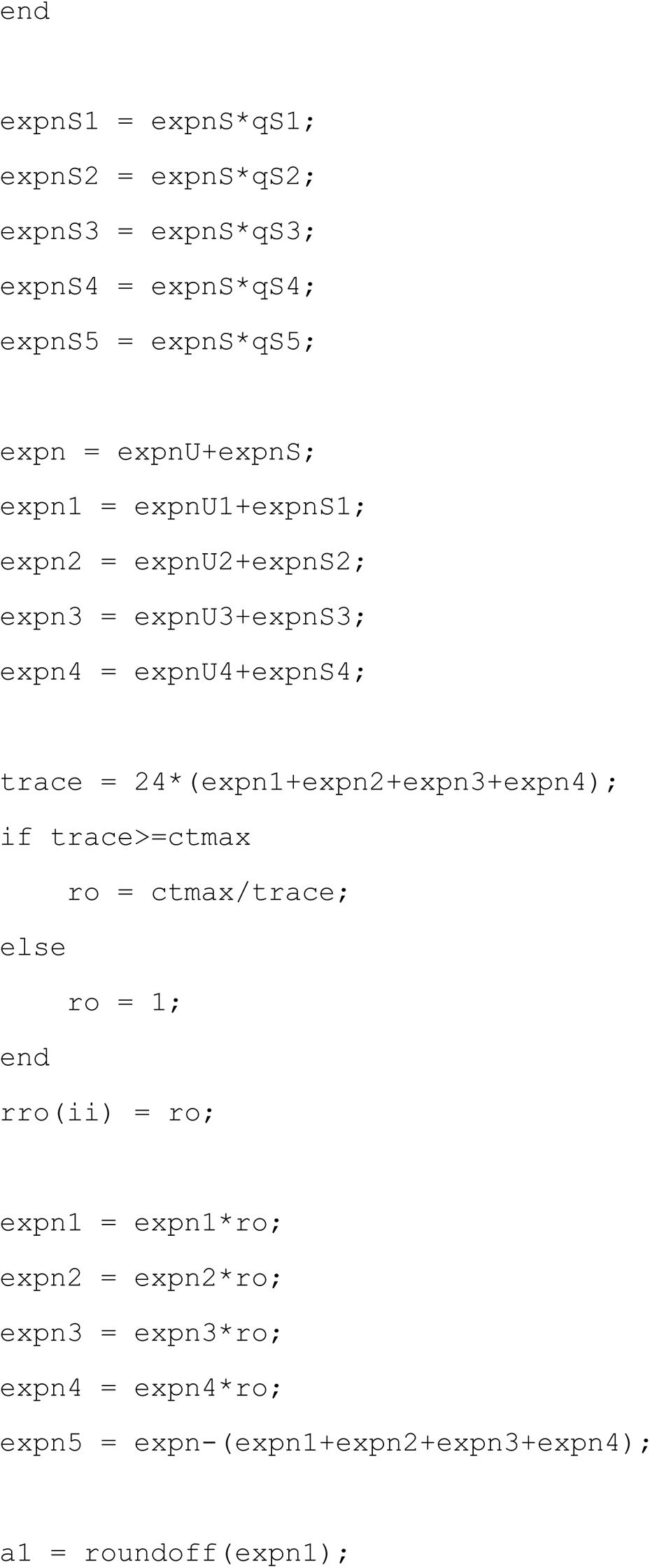

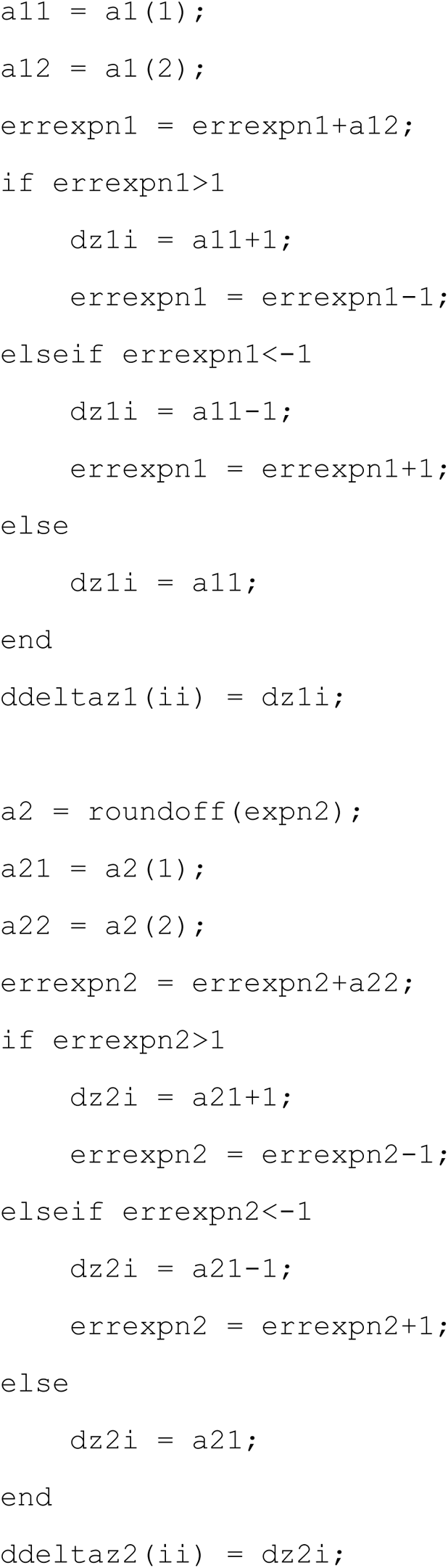

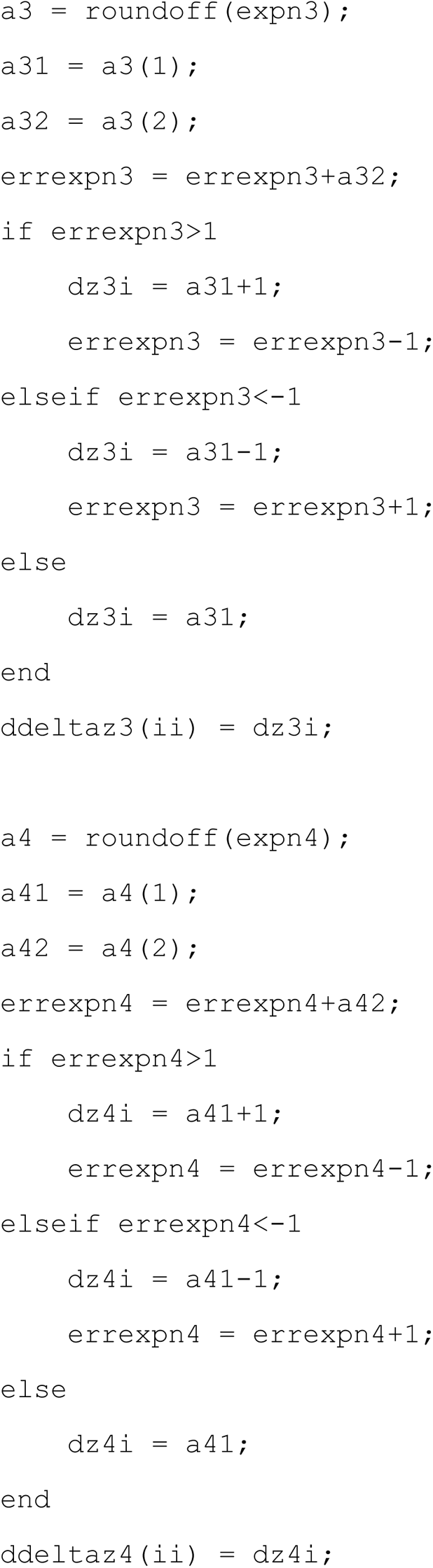

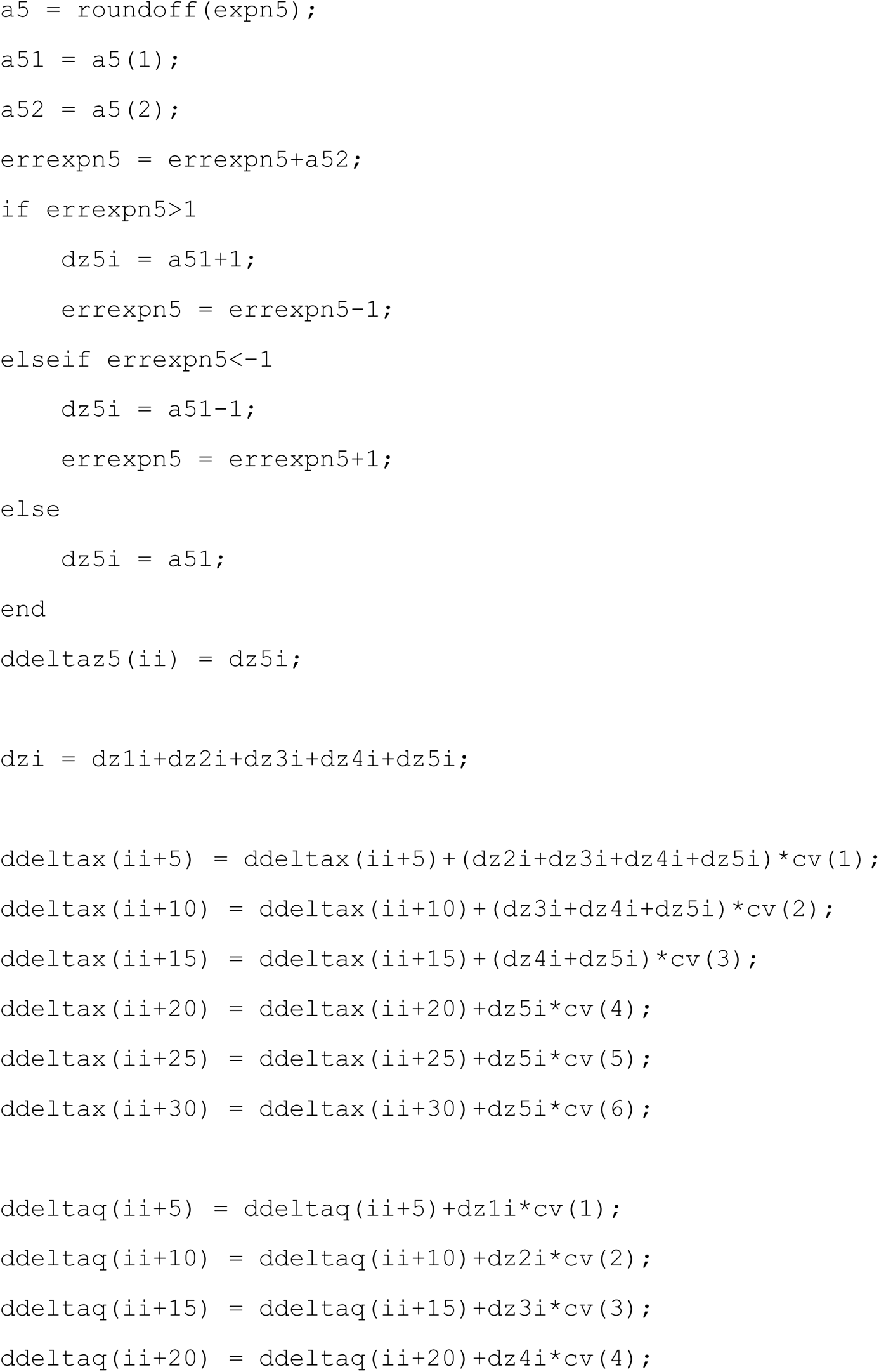

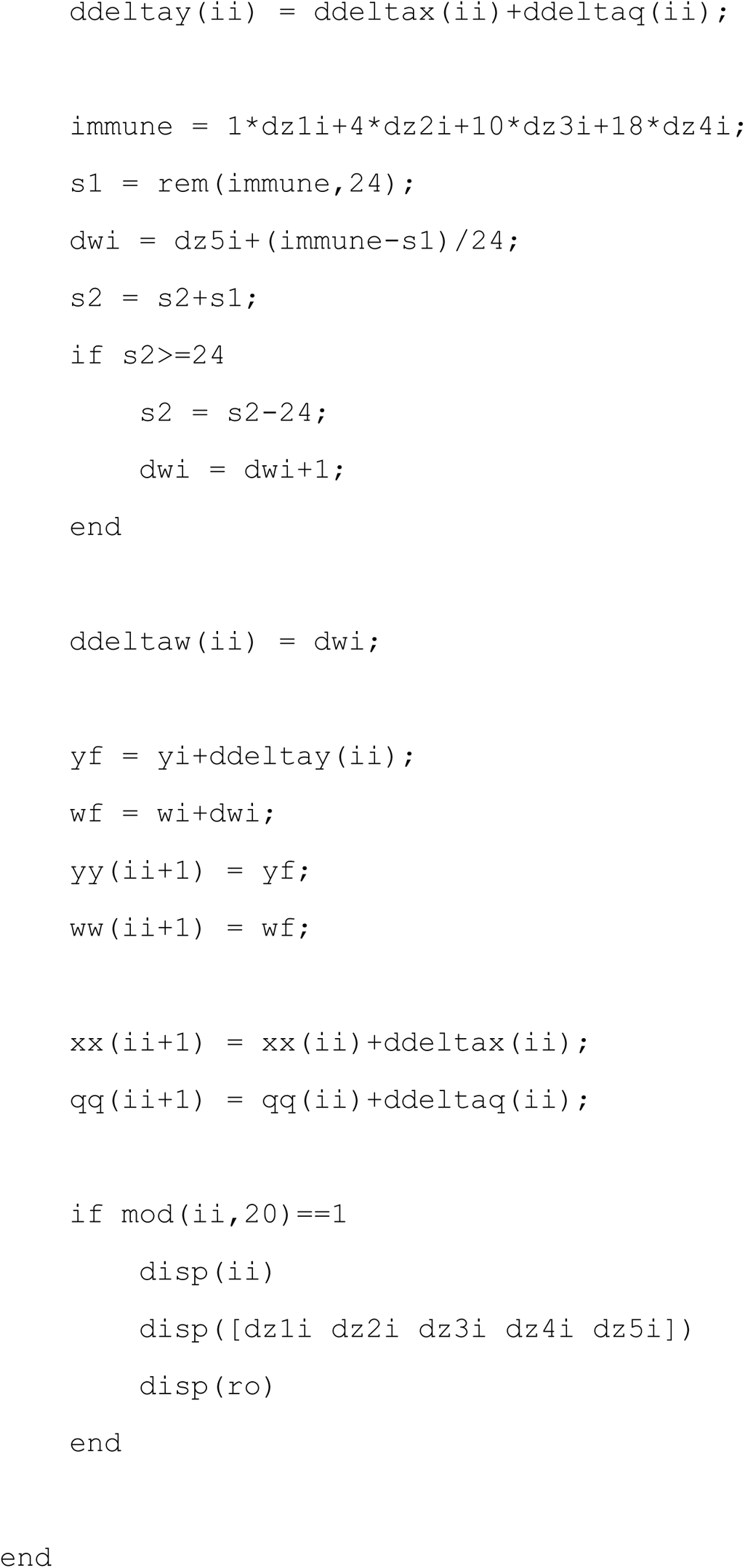

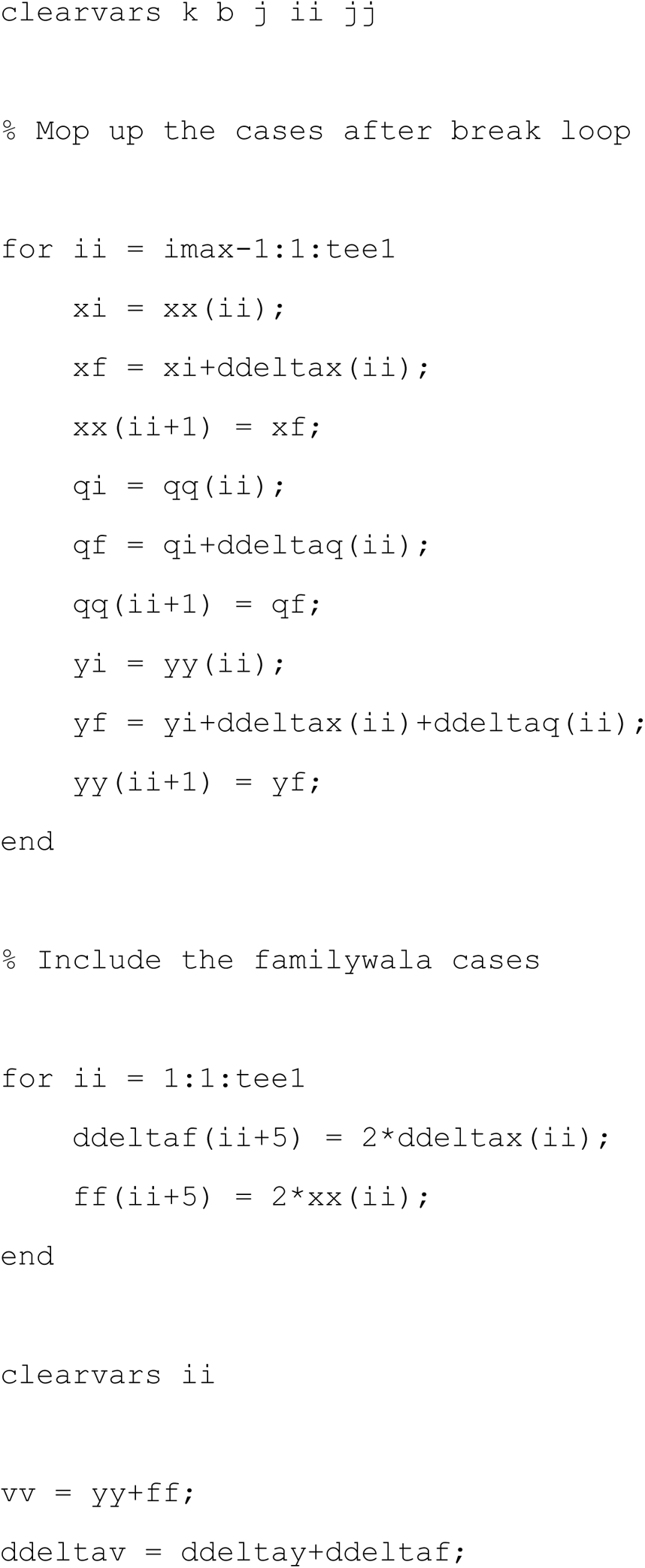

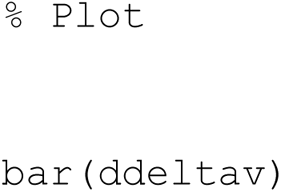

We have verified that after performing these steps, Fig. 1 appears on the screen minus the beautification.

